# Estimating severity and rate of change of depressive symptoms in adolescence: a comparison of functional principal component analysis and mixed effects models

**DOI:** 10.64898/2026.04.09.26350500

**Authors:** María A Hernandez, Alex SF Kwong, Cai Li, Andrew J Simpkin, Robyn E Wootton, Carol Joinson, Ahmed Elhakeem

## Abstract

Understanding depressive symptoms dynamics and their determinants is crucial for designing effective mental health support initiatives. This study compared two methods for describing youth depressive symptoms trajectories and investigated associations of early-life factors (maternal education, maternal perinatal depression, domestic violence, physical, emotional, or sexual abuse, bullying victimisation, psychiatric disorder) with trajectory features. Prospective data from 8,264 mostly White European participants (54% female), including self-reported Short Moods and Feelings Questionnaires on ten occasions between 10-25 years, were used. Trajectories were summarised using functional principal component analysis (FPCA) and P-splines linear mixed-effect (PLME) models. Estimated derivatives were used to obtain magnitude and age of peak symptoms and peak symptoms velocity. Both methods performed comparably, but PLME models tended to over-smooth trajectories. Peak symptoms and peak velocity were higher and occurred >1 year earlier in females than males. All early-life factors were associated with higher peak symptoms, and most associated with higher and earlier peak velocity. Abuse and bullying additionally associated with earlier age of peak symptoms. FPCA is a useful alternative for characterising depressive symptoms trajectories and informing time-sensitive preventative measures to reduce impact of depression before symptoms reach their peak. Early-life stressors may accelerate timeline and intensity of symptoms escalation during adolescence.

**Lay summary:** Understanding development of depressive symptoms and factors shaping them is crucial for designing effective mental health support initiatives. This study used data from over 8,000 young people regularly followed up from before birth to compare two cutting-edge methods for describing depressive symptoms trajectories and examined how known risk factors for adulthood depression relate to the severity and rate of change of depressive symptoms in adolescence. We found that both methods performed well and that the peaks in depressive symptoms and their rate of change were, on average, higher and occurred over a year earlier in females than males. Our findings additionally suggest that early-life stressors (e.g., abuse, bullying) may accelerate the development of depression, highlighting the importance of early prevention.

## INTRODUCTION

Major depressive disorder is a common illness and a leading cause of global disease burden (GBD 2023 Disease and Injury and Risk Factor Collaborators, 2025), with adolescence a critical developmental window for its onset (McGrath et al., 2023).

Both adolescent-onset depressive disorder (Johnson et al., 2018, Koenen et al., 2014) and elevated subclinical symptoms during this period (Portogallo et al., 2024) are key predictors for major depressive disorder in adulthood. Examining quantitative (i.e., continuous) symptoms trajectories in a general population, instead of binary diagnostic thresholds, is particularly valuable during adolescence as subclinical symptoms are common and increasing in this period (Bertha and Balázs, 2013), and associated with elevated risk of depression in adulthood (Fergusson et al., 2005).

Understanding longitudinal nature of these symptoms requires sophisticated analyses of repeated measurements to identify adverse patterns in symptoms trajectories. This can include determining the magnitude and age of peak depressive symptoms (i.e., the trajectory maxima) and the magnitude and age of peak symptom velocity, which represents the fastest rate of symptom change (Grimm et al., 2011). Studying these trajectory features and their early-life determinants can inform the timing of preventative interventions to improving life course trajectories and prevent more severe depression outcomes (Kwong et al., 2019a).

Mixed effects models (Laird and Ware, 1982) and functional data analysis (Ramsay, 1982) are two prominent but conceptually distinct approaches to trajectory analysis. Both estimate the population-average trajectory and individual-specific deviations, however, while mixed effect models treat data as discrete repeated measurements, functional data analysis views observations as realisations of underlying continuous functions. Mixed effects models are commonly used in mental health literature, including for depressive symptoms trajectories (Kwong et al., 2019a, Edmondson-Stait et al., 2025, Motoc et al., 2025, Rawana and Morgan, 2014). Functional data analysis has been used on various trajectory outcomes including physical growth (Feng et al., 2020, Karuppusami et al., 2022, Simpkin et al., 2017), biomarkers (Ruffieux et al., 2023), and cognitive decline (Ségalas et al., 2024). However, to the best of our knowledge, it has not yet been applied to depressive symptoms.

Consequently, its comparative performance against mixed effects models for characterising youth depressive symptoms trajectories remains unknown. Beyond trajectory characterisation, examining early life determinants of trajectory features can pinpoint critical developmental windows where stressors accelerate symptom escalation. Such insights are essential for timing preventative interventions to reduce symptom velocity and magnitude of the adolescent depressive peak.

The aim of this study was to (i) compare functional data analysis with mixed effects models for describing depressive symptoms trajectories throughout adolescence, and (ii) investigate associations between known early life risk factors for adulthood depression and key features estimated from the fitted trajectories, specifically the magnitude and age of peak symptoms, and the magnitude and peak symptoms velocity.

## METHODS

### Avon Longitudinal Study of Parents and Children (ALSPAC)

ALSPAC is a prospective birth cohort study that recruited pregnant women resident in Avon, UK with an expected date of delivery between April 1991 and December 1992 (Boyd et al., 2013, Fraser et al., 2013, Northstone et al., 2019). The initial number of pregnancies enrolled was 14,541. Of these initial pregnancies, there was a total of 14,676 fetuses, resulting in 14,062 live births and 13,988 children who were alive at 1 year of age. When the children were around 7 years of age, an attempt was made to bolster the initial sample with eligible cases who had failed to join the study originally. The total sample size for analyses using any data collected after age 7 years was 15,447 pregnancies, resulting in 15,658 fetuses. Of these 14,958 children were alive at 1 year of age. Data were collected from the parents and their children using questionnaires, data extraction from medical records, health records linkage, and research clinic assessments up to the last completed contact. Study data were collected and managed using Research Electronic Data Capture (REDCap) tools hosted at University of Bristol (Harris et al., 2009). REDCap is a secure, web-based software platform designed to support data capture for research studies.

### Longitudinal assessment of depressive symptoms

Depressive symptoms were self-reported between 2002-2018 across ten time points from mean age 10.6 to 25.8 years using the Short Mood and Feelings Questionnaire (SMFQ), which is a validated instrument for measuring depressive symptoms (Angold et al., 1995, Kwong, 2019, Turner et al., 2014, Eyre et al., 2021). At each age, presence of depressive symptoms during the previous two weeks was recorded by the participant responses to 13 questions (**Appendix S1**). Responses for each question were reported on a 3-point scale (0 = not true, 1 = sometimes, 2 = true). Responses were summed to create a total score ranging from 0 to 26, with higher scores indicating greater depressive symptoms (Angold et al., 1995).

### Early life risk factors

Low childhood socioeconomic position (SEP) (Damianakou et al., 2024), perinatal maternal depression (Tirumalaraju et al., 2020), childhood trauma (Mandelli et al., 2015), and psychiatric disorder (Mulraney et al., 2021, Erskine et al., 2016) are known early life risk factors for adult depression and were selected to examine their association with peak symptoms outcomes.

Details of risk factor assessment is provided in **Appendix S1**. Briefly, SEP was defined by self-reported maternal education in pregnancy (Galobardes et al., 2006) and was categorised according to the International Standard Classification of Education 97/2011 as high, medium, or low (Cadman et al., 2023). Perinatal maternal depression was assessed via the Edinburgh Postnatal Depression Scale (score >12) during pregnancy and 8 months postpartum, and was coded as any depression vs. none (Paul and Pearson, 2020, Hermans et al., 2026). Childhood trauma was assessed by parent and participants reports between 0-10 years and supplemented by retrospective participant questionnaires at age 22 years (Croft et al., 2019). The responses were used to derive three binary variables: domestic violence, physical, emotional, and sexual abuse, and peer bullying victimisation up to 10 years of age. Childhood psychiatric diagnoses (attention deficit/hyperactivity, anxiety, depressive, oppositional-conduct, and pervasive development disorders) were identified using parent-reported Development and Well-Being Assessment at age 8 years (Goodman et al., 2000) and coded as a binary variable comparing children with and without any disorder.

### Statistical analysis

#### Trajectory analysis and peak estimation

Depressive symptoms trajectories were analysed using two approaches based on functional principal component analysis (FPCA) (Besse and Ramsay, 1986) and linear mixed effects (LME) models (Laird and Ware, 1982). FPCA is a functional data analysis approach, conceptually aligned with unsupervised learning, that treats repeated measurements as discrete observations of an underlying continuous smooth trajectory over time. FPCA estimates the population-average trajectory and then identifies the principal sources of variation where individual trajectories deviate from this average. These patterns of variation are summarised by functional principal components (FPCs). Formally, individual trajectories are represented using the Karhunen-Loève expansion, which expresses each individual’s trajectory as the mean function plus a weighted combination of these FPCs. These weights, known as FPC scores, quantify how an individual’s trajectory deviates from a population-average trajectory. In contrast, the LME framework models the repeated outcome as a linear combination of fixed effects (representing the population-average trajectory) and random effects (capturing individual-specific deviations from the average trajectory).

To accommodate nonlinear trajectories, both FPCA and LME models can be extended to use connected piecewise polynomials called basis splines (B-splines). In FPCA, splines are used for smoothing the mean and covariance functions, whereas in LME models, they allow the fixed and random effects to be represented as smooth functions of age. In this way, random effects coefficients in LME models play a role analogous to FPC scores in FPCA: they both quantify how individual trajectories differ from the mean function (**Figure S4**). In this study, smoothing was implemented using cubic P-splines (penalised B-splines), which combine a B-spline basis with a difference-based penalty that is controlled by a smoothing parameter to prevent overfitting (Eilers and Marx, 1996).

Analyses were done separately in males and females with two or more SMFQ measurements. Functional model components for FPCA were estimated using the fast covariance estimation algorithm (Xiao et al., 2018), which uses tensor-product bivariate P-splines for covariance estimation. The smoothing parameter was selected via leave-one-subject-out cross-validation and FPC scores were subsequently obtained as conditional expectations based on principal components analysis (Yao et al., 2005). In the P-spline LME (PLME) model, the population-average trajectory and individual-specific deviations were estimated using fixed and random P-splines, respectively (Hernandez et al., 2025, Djeundje and Currie, 2010) and smoothing parameter was selected using restricted maximum likelihood estimation. FPCA and PLME analyses were implemented using the face (Li C, 2025) and psme (Li Z, 2025) R libraries, respectively. A detailed description of the FPCA and PLME methods is provided in **Appendix S1**.

FPCA and PLME fitted trajectories were obtained over an age grid of 200 equidistant points from ages 10-25 years (equivalent to 1-month intervals across the observed age range). Peak features of adolescent symptom trajectories were then estimated from these individual-level fitted trajectories up to age 20 years. The magnitude and age of peak depressive symptoms were identified by locating the highest local maximum on each fitted trajectory based on the derivative sign change method with an extra quadratic refinement procedure (**Appendix S1**). Symptom velocity curves were obtained using cubic smoothing spline differentiation of each individual’s fitted trajectory at each point on the age grid and the same sign change procedure was then used to identify peak symptoms velocity (rate of most rapid increase in depressive symptoms: score/year) and the corresponding age at peak velocity (in years).

#### Comparison of FPCA and PLME trajectories and peak estimates

Model performance of FPCA and PLME was assessed by using the root mean square error (RMSE) to quantify the difference between observed and fitted trajectories. Plots of the model residuals against age were generated and used to evaluate whether residual variability was constant across the age range. Trajectory recovery performance was assessed by inspecting the estimated mean and individual-specific score trajectories for all participants and the estimated trajectories against observed scores for randomly selected participants. We assessed efficiency by comparing the computation time for each method. Agreement between the FPCA and PLME based estimates of magnitude and age of peak symptoms, and magnitude and age of peak velocity was evaluated using Pearson correlation and Bland-Altman analyses. For Bland-Altman analysis, we plotted the individual-level mean and difference scores and used the mean difference and 95% limits of agreement to assess systematic bias and consistency of agreement between the methods.

#### Association of early life risk factors with peak symptoms and peak velocity

We used linear regression models to examine the associations of sex and each early life risk factor with our estimated magnitude and age of peak symptoms and peak velocity from trajectory analyses. To compare methods, analyses were done for outcomes estimated from FPCA and PLME, as well as an average of the FPCA-PLME estimates. For clarity of presentation, only results for the FPCA estimates are shown in the main paper (selected because of the larger sample size) with other results presented in the supplement. Risk factor models were fitted in males and females combined (to increase statistical power) with adjustment made for sex and maternal education, using all available data for each risk factor in participants with data on outcomes. We examined sex differences in associations by fitting additional models with an interaction term between each risk factor and sex. Where evidence of a sex interaction was found, we calculated marginal linear predictions to obtain estimates separately for males and females. All analyses were performed in R 4.5.0 (R Project for Statistical Computing).

## RESULTS

### Participant characteristics

A total of 3,800 males and 4,464 females had more than one SMFQ measurement and were included in depressive symptoms trajectory analysis (**Table 1**). The total number of SMFQ observations was 19,416 for males and 27,346 for females; the median (interquartile range) number of observations per study participant was 4 (4) in males and 6 (5) in females. When compared with the included participants, those excluded from the trajectory analysis (due to having fewer than two (or no) SMFQ observations) had a higher proportion of males, higher prevalences of low maternal education, prenatal maternal depression, domestic violence, and psychiatric disorder, slightly lower prevalence of bullying victimisation, and similar levels of physical, emotional, or sexual abuse (**Table S1**).

**Table 1.** Descriptive statistics of the Short Mood and Feelings Questionnaire (SMFQ) in the trajectory analysis sample.

| Occasion | Males, N=3800 |  |  | Females, N=4464 |  |  |
| --- | --- | --- | --- | --- | --- | --- |
|  | N participants | Age, mean (SD) | SMFQ, mean (SD) | N participants | Age, mean (SD) | SMFQ, mean (SD) |
| 1 | 3235 | 10.6 (0.3) | 4.2 (3.5) | 3536 | 10.6 (0.3) | 3.9 (3.5) |
| 2 | 3200 | 12.8 (0.2) | 3.6 (3.4) | 3377 | 12.8 (0.2) | 4.3 (4.2) |
| 3 | 2904 | 13.8 (0.2) | 4.1 (3.8) | 3051 | 13.8 (0.2) | 5.7 (4.9) |
| 4 | 1947 | 16.7 (0.3) | 4.3 (4.6) | 2877 | 16.7 (0.2) | 7.0 (6.0) |
| 5 | 1826 | 17.8 (0.4) | 5.6 (4.8) | 2512 | 17.8 (0.4) | 7.3 (5.5) |
| 6 | 1162 | 18.7 (0.5) | 5.3 (5.0) | 2126 | 18.6 (0.5) | 7.6 (6.2) |
| 7 | 1160 | 22.0 (0.5) | 4.9 (4.8) | 2120 | 22.0 (0.5) | 6.1 (5.9) |
| 8 | 1331 | 22.9 (0.5) | 5.4 (4.9) | 2548 | 22.9 (0.5) | 6.6 (5.8) |
| 9 | 1336 | 23.9 (0.5) | 6.2 (5.6) | 2593 | 23.9 (0.5) | 7.5 (6.2) |
| 10 | 1305 | 25.8 (0.5) | 5.5 (5.5) | 2606 | 25.8 (0.5) | 7.5 (6.7) |

Table shows the number of participants, age, and SMFQ depressive symptoms score at each of the 10 SMFQ occasions in the participants included in the trajectory analysis (i.e., those with two or more SMFQ occasions).

### Depressive symptoms trajectories

FPCA and PLME depressive symptoms score trajectories are presented in **Figure 1**. Both methods showed that females had higher and steeper mean symptom score trajectories. The trajectory pattern was consistent across both methods, but the PLME estimated trajectories were smoother than the FPCA trajectories, particularly for males. Inspecting the individual-specific trajectories also showed smoother curves from PLME, with the FPCA trajectories displaying more peaks (**Figure 1**).

**Figure 1.**
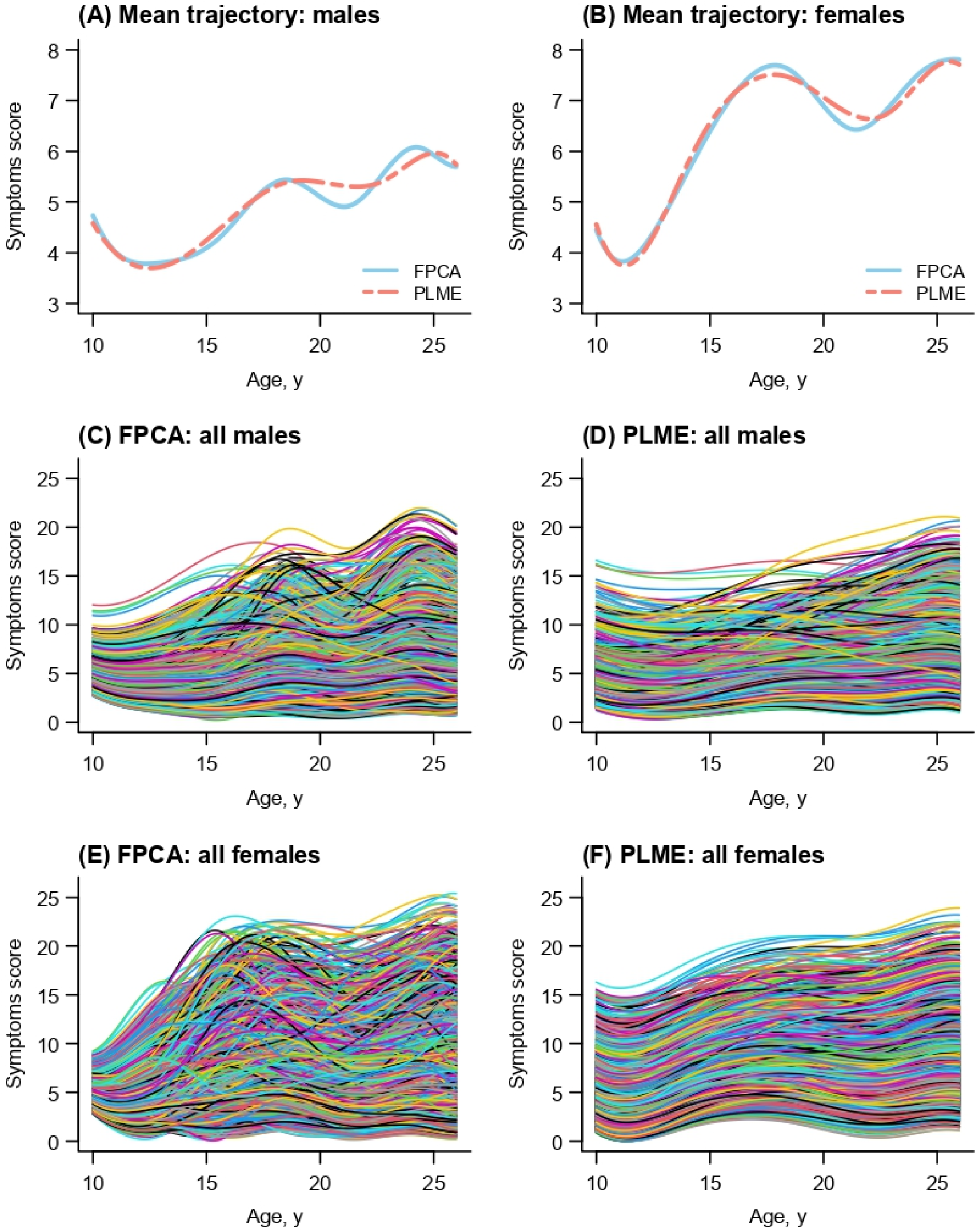
Estimated depressive symptoms trajectories. Figure shows the FPCA and PLME estimated population-average SMFQ depressive symptoms score trajectories in males and females (A, B), and the FPCA and LME predicted individual-specific trajectories in males (C, D) and females (E, F).

Symptoms velocity trajectories showed an even more exaggerated difference in the degree of smoothing between the PLME and FPCA curves (**Figure S1**).

Both methods demonstrated good overall goodness-of-fit in modelling depressive symptoms via RMSE, though accuracy was higher for males than for females across both methods. For males, FPCA and PLME models performed similarly: FPCA achieved an RMSE of 2.2 (in units of *SMFQ score)* and PLME had an RMSE of 2.3. For females, FPCA showed better accuracy (RMSE = 2.4) than PLME (RMSE = 3.1). Plots of the model residuals supported this by showing a wider range for the PLME than FPCA residuals (**Figure S2**). Examining estimated SMFQ score trajectories against observed values further demonstrated that both methods performed well (**Figure 2**). Total estimation time was 18 *minutes* for FPCA and 23 *seconds* for PLME (using an intel core i7 16GB RAM Dell laptop).

**Figure 2.**
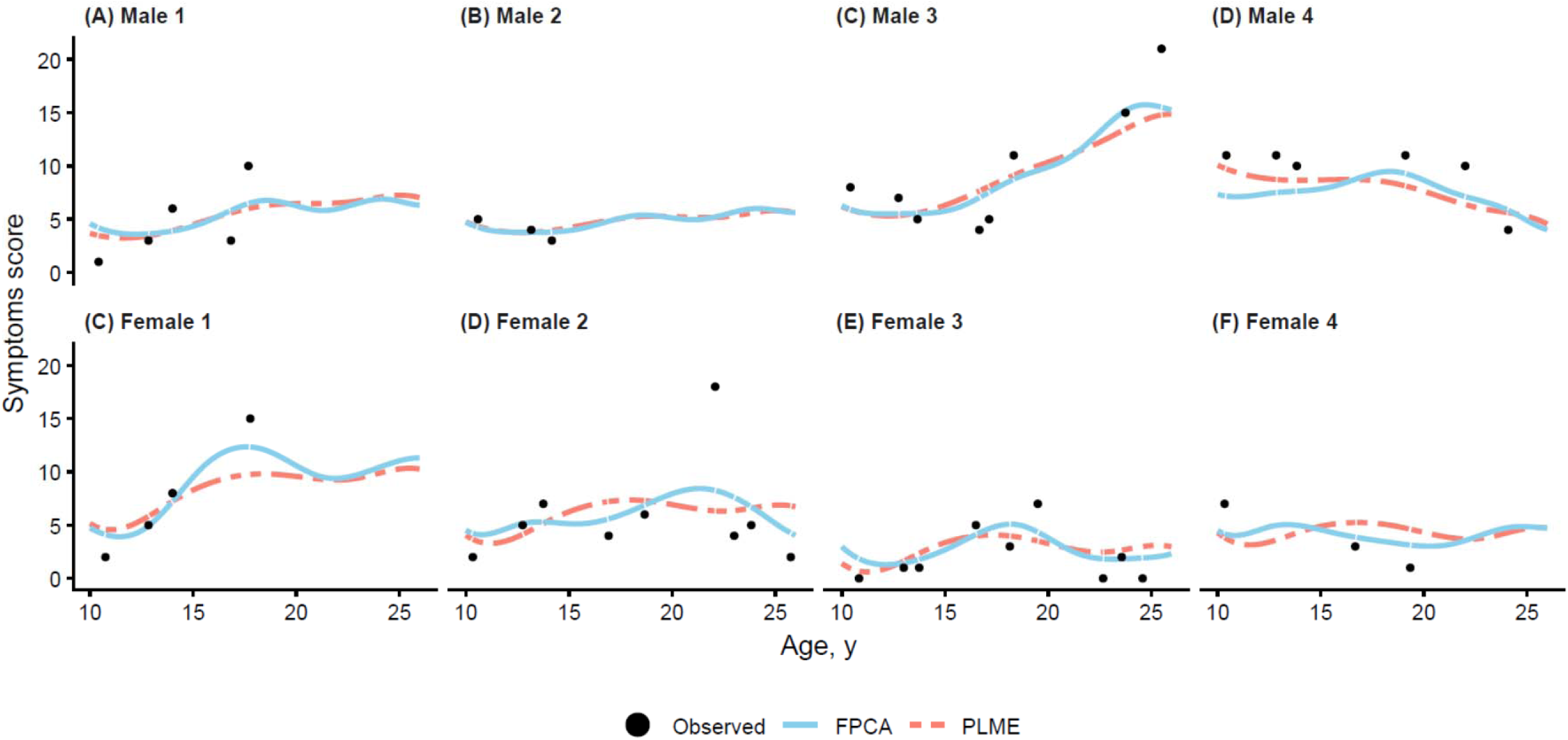
Observed values versus estimated depressive symptoms trajectories for eight randomly selected participants. Figure shows the FPCA and PLME estimated SMFQ depressive symptoms score trajectories versus the observed scores for four randomly selected males (A-D) and four randomly selected females (C-F).

### Magnitude and age of peak symptoms and peak velocity

Of the 8,264 participants included in the trajectory analyses, peak velocity (and age at peak velocity) was estimated for 8,254 participants (both FPCA and PLME) and peak symptoms (and age at peak symptoms) was estimated for 7,546 (FPCA) and 6,446 (PLME) participants. **Figure 3** summarises these estimates in 6,137 participants (3,905 females) with all four pairs of peak symptoms/velocity estimates. Both methods showed that females had higher mean peak symptoms and higher mean peak velocity than males. Both methods also showed that females had younger mean age at peak symptoms and younger age at peak velocity than males. There was more variability between individuals, and a narrower range of values, for estimates from FPCA than those from PLME, particularly for the age at peak velocity. For example, the standard deviation (SD) around the mean age at peak velocity was 1.1 and 0.4 *years* for FPCA and PLME in males, and the corresponding SD estimates in females were 1.8 and 0.1 *years* (**Table S2**). Findings were similar when looking at each unique sample (**Table S2**).

**Figure 3.**
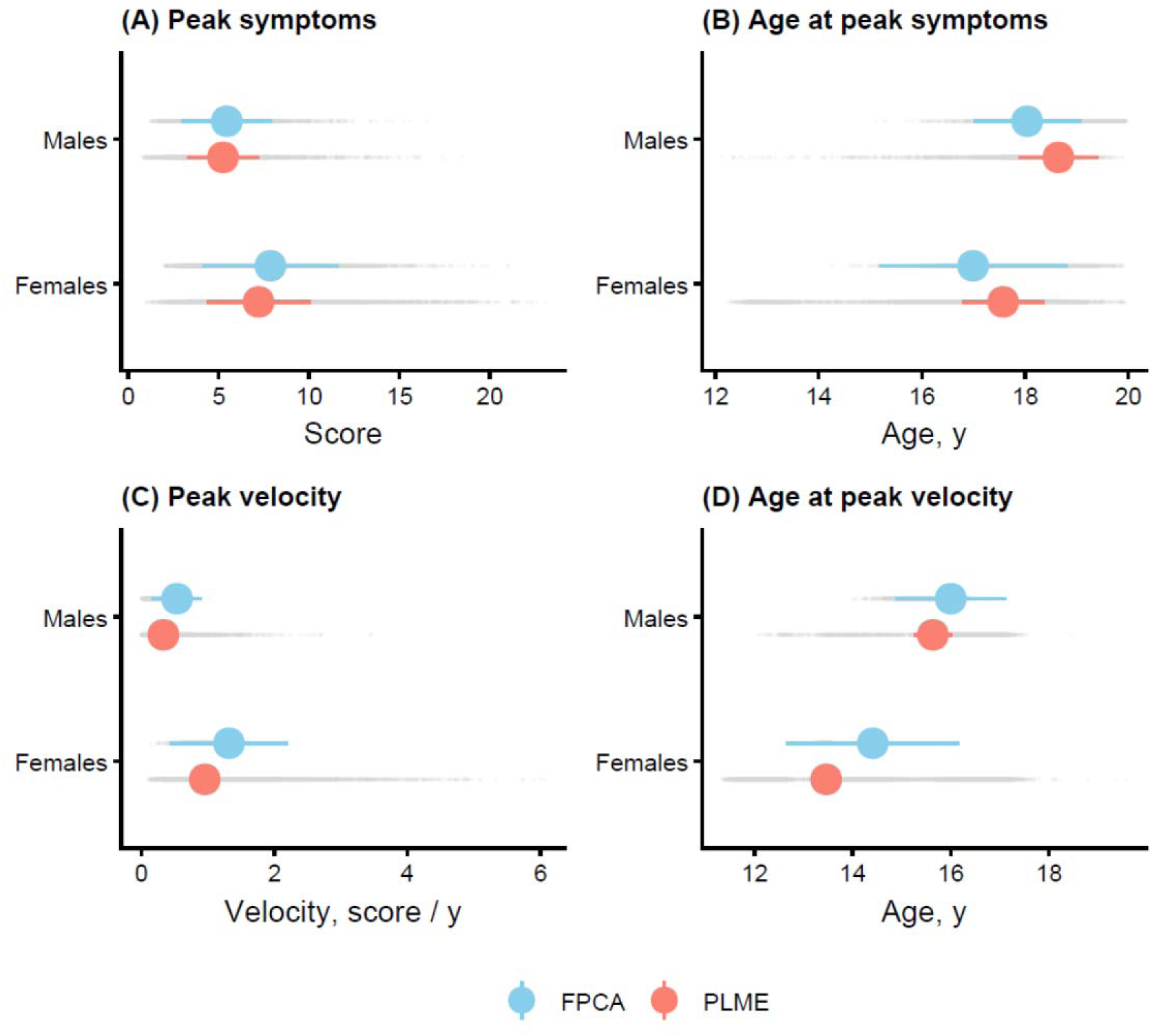
Magnitude and age of peak symptoms and peak symptoms velocity. Figure summarises the FPCA and PLME based estimates of the magnitude (A) and age (B) of peak depressive symptoms score, and the magnitude (C) and age (D) of peak symptoms velocity in males and females. The coloured points are the mean values, and the coloured horizontal bars represent the mean +/− 1 SD. The grey points are the values for each study participant. The magnitude and age of peak symptoms and peak symptoms velocity was estimated for 7,506 and 8,254 participants after FPCA analysis and for 6,446 and 8,254 participants after PLME analysis, respectively. The data in this figure are based on 6,137 participants with estimates from both methods (n=2,232 males, n=3,905 females). The numerical estimates in this figure and the estimates for the available samples for each method are provided in Table S2.

There were mostly negligible or weak associations between the magnitude and age of peak symptoms in males and females, including for either FPCA or PLME estimates (**Table S3**). There was virtually no association between FPCA based estimates of the magnitude and age of peak velocity in either males or females, while the PLME based estimates showed weak positive (females) and negative (males) associations (**Table S3**).

Results of analysis of agreement between FPCA and PLME peak estimates are presented as correlation coefficients (**Table S3**) and Bland-Altman plots (**Figure S3**).

Reassuringly there were positive associations between all pairs of estimates, though with differences in strength. There were very strong associations between peak symptoms estimates, and mostly moderate associations for other estimates (i.e., age at peak symptoms, peak velocity, and age at peak velocity), for both males and females (**Table S3**).

Bland-Altman analysis further supports the agreement between FPCA and PLME across the various peak outcomes. For peak symptoms, FPCA tended to yield higher estimates than PLME, with difference increasing with symptom severity, particularly in females (**Figure S3**). Similarly for peak velocity, FPCA systematically produced higher velocity estimates than PLME, with differences increasing for larger peak velocities. With respect to age at peak symptoms, the bias was small indicating good concordance between methods. However, for age at peak velocity, a linear trend in the differences emerged, suggesting that the level of agreement depends on the magnitude of the age. While visual inspection suggests that agreement for the age at peak symptoms remained consistent across the observed range, a proportional bias was evident for both peak symptom velocity and the age at peak velocity. Despite these trends, most observations fell within the 95% limits of agreement, although these limits were notably wider for females than males across most parameters.

### Association of early life factors with peak symptoms and peak velocity

Characteristics of the participants included for examining associations of sex and early life risk factors with FPCA based magnitude and age of peak symptoms and peak velocity are presented in **Table 2**. The prevalence of risk factors in the sample varied from 6.2% for any psychiatric disorder at age 8 years to 24.5% for any physical, emotional, or sexual abuse up to age 10 years.

**Table 2.**
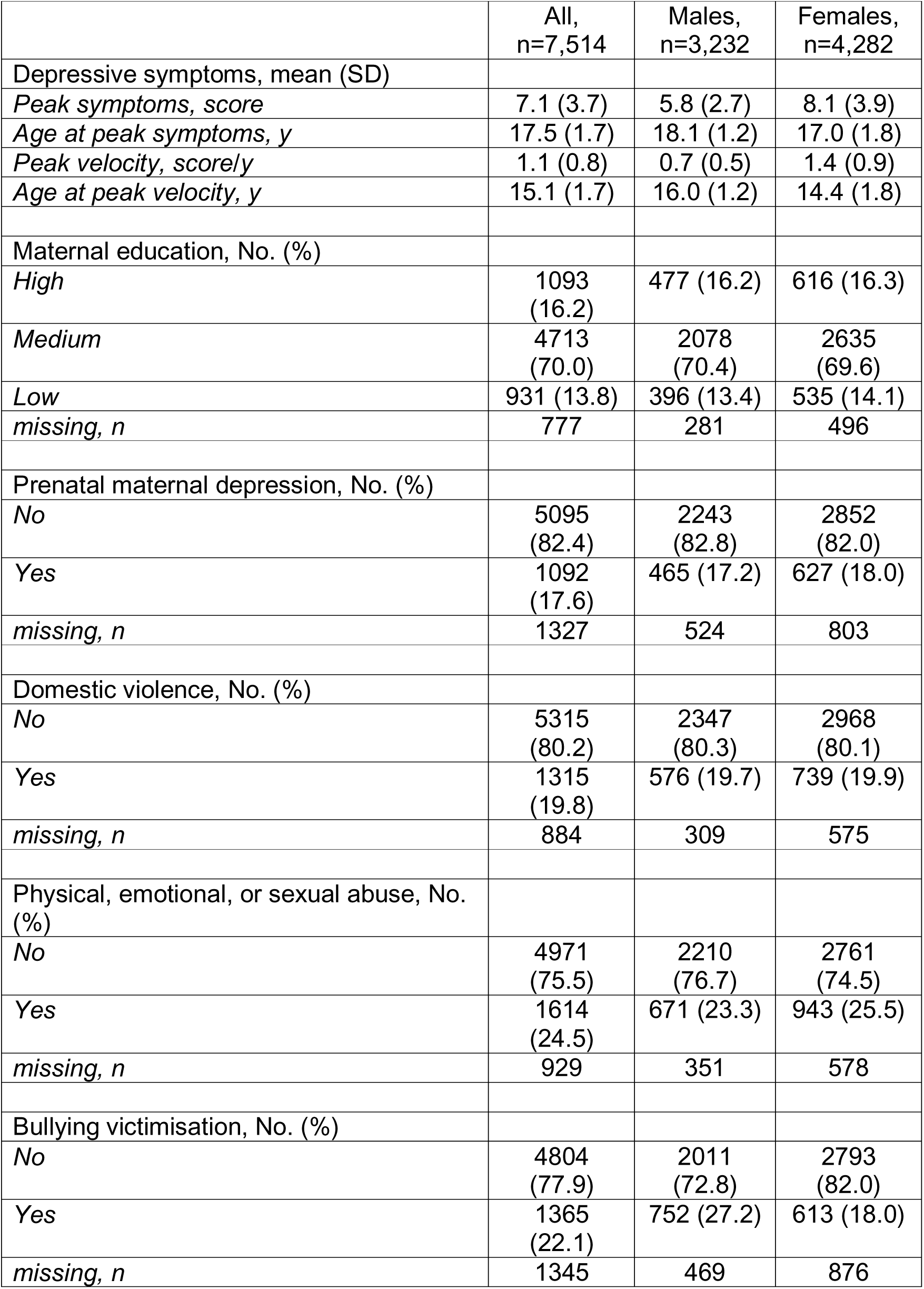
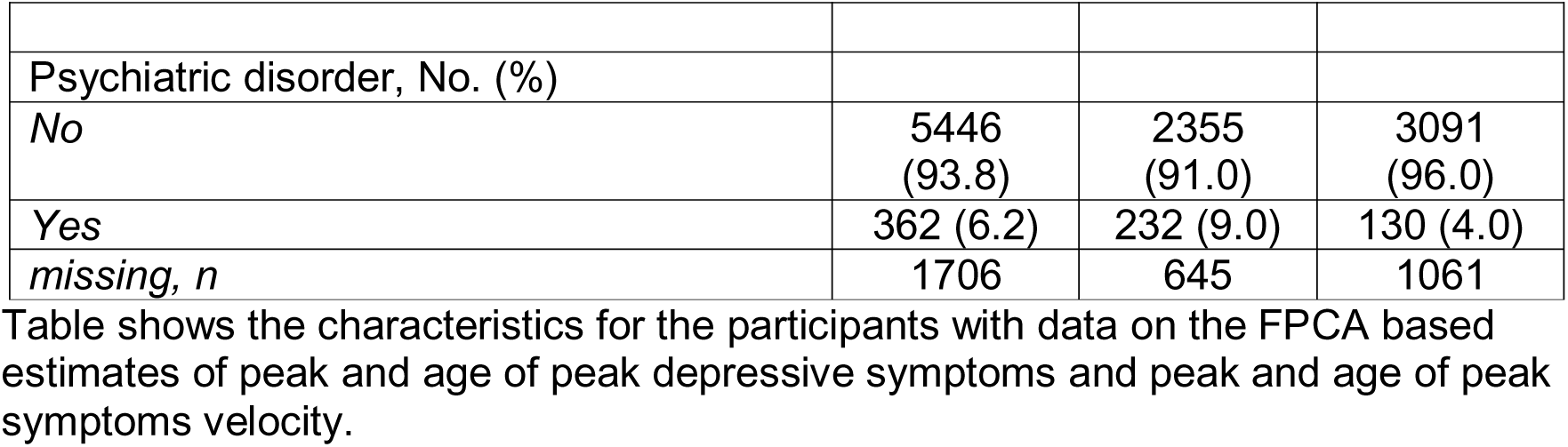
Characteristics of the participants included for examining associations between early life factors and FPCA based depressive symptoms outcomes.

**Table 3** presents the mean differences in depressive symptoms outcomes for sex and each early life risk factor. Analysis of the sex differences in peak symptoms and peak symptoms velocity included 7.510 participants. When compared with males, females had higher peak symptoms (mean difference in SMFQ score: 2.4, 95%CI: 2.2 to 2.5), higher peak velocity (0.74 *score/year*, 0.70 to 0.77), younger age at peak symptoms (-1.1 *years*, -1.2 to -1.0) and younger age at peak velocity (-1.6 *years*, - 1.7 to -1.5).

**Table 3.** Association of early life factors with FPCA based magnitude and age of peak symptoms and peak symptoms velocity.

|  | N | Mean difference (95 % CI) |  |  |  |
| --- | --- | --- | --- | --- | --- |
|  |  | Peak symptoms |  | Peak symptoms velocity |  |
|  |  | Peak symptoms (SMFQ score) | Age at peak symptoms (y) | Peak velocity (SMFQ score / y) | Age at peak velocity (y) |
| Sex |  |  |  |  |  |
| *Males* | 3232 | 0 (ref) | 0 (ref) | 0 (ref) | 0 (ref) |
| *Females* | 4282 | 2.36 (2.21 to 2.52) | -1.10 (-1.17 to -1.02) | 0.74 (0.70 to 0.77) | -1.59 (-1.66 to -1.52) |
| Maternal education<sup>a</sup> |  |  |  |  |  |
| *High* | 1093 | 0 (ref) | 0 (ref) | 0 (ref) | 0 (ref) |
| *Medium* | 4713 | 0.26 (0.03 to 0.49) | 0.17 (0.06 to 0.27) | 0.06 (0.01 to 0.11) | 0.09 (-0.01 to 0.19) |
| *Low* | 931 | 0.55 (0.25 to 0.85) | 0.31 (0.17 to 0.45) | 0.09 (0.02 to 0.15) | 0.06 (-0.07 to 0.20) |
| Perinatal maternal depression<sup>b</sup> |  |  |  |  |  |
| *No* | 5003 | 0 (ref) | 0 (ref) | 0 (ref) | 0 (ref) |
| *Yes* | 1048 | 1.13 (0.90 to 1.36) | 0.04 (-0.07 to 0.15) | 0.14 (0.08 to 0.19) | -0.16 (-0.27 to -0.06) |
| Domestic violence<sup>b</sup> |  |  |  |  |  |
| *No* | 5174 | 0 (ref) | 0 (ref) | 0 (ref) | 0 (ref) |
| *Yes* | 1258 | 0.63 (0.42 to 0.85) | -0.03 (-0.13 to 0.07) | 0.04 (0.00 to 0.09) | -0.18 (-0.27 to -0.08) |
| Physical, emotional, or sexual abuse<sup>b</sup> |  |  |  |  |  |
| *No* | 4838 | 0 (ref) | 0 (ref) | 0 (ref) | 0 (ref) |
| *Yes* | 1544 | 1.28 (1.09 to 1.48) | -0.12 (-0.22 to -0.03) | 0.19 (0.15 to 0.23) | -0.23 (-0.32 to -0.14) |
| Bullying victimisation<sup>b</sup> |  |  |  |  |  |
| No | 4693 | 0 (ref) | 0 (ref) | 0 (ref) | 0 (ref) |
| Yes | 1317 | 1.19 (0.98 to 1.4) | -0.23 (-0.33 to -0.13) | 0.14 (0.1 to 0.19) | -0.33 (-0.42 to -0.23) |
| Psychiatric disorder<sup>b</sup> |  |  |  |  |  |
| No | 5293 | 0 (ref) | 0 (ref) | 0 (ref) | 0 (ref) |
| Yes | 351 | 1.28 (0.90 to 1.66) | -0.08 (-0.26 to 0.10) | 0.13 (0.05 to 0.22) | -0.28 (-0.44 to -0.11) |

Results were obtained from separate linear regression models. <sup>a</sup> adjusted for sex. <sup>b</sup> adjusted for sex and maternal education.

Up to 6,737 (55% female) participants were included for examining associations between early life risk factors and peak outcomes. All risk factors (i.e., lower maternal education, perinatal maternal depression, domestic violence, physical, emotional, or sexual abuse, bullying victimisation, and psychiatric disorder) were associated with higher peak symptoms and higher peak velocity (**Table 3**). For example, mean difference in peak symptoms between those with (vs. without) a psychiatric disorder at age 8 years was 1.28 *SMFQ score points* (95%CI: 0.90 to 1.66). Bullying victimisation and abuse were also associated with a younger age of peak symptoms (e.g., mean difference in age at peak symptoms for any peer bullying victimisation up to age 10 years (vs. none) was -0.23 *years*, -0.33 to -0.13), whereas lower maternal education was associated with *older* age of peak symptoms. All risk factors, apart from maternal education, were/associated with a younger age at peak velocity. Associations were mostly consistent across FPCA and PLME based estimates, and when using an average of the two estimates (**Table S4**).

The observed associations between lower maternal education, perinatal maternal depression, bullying victimisation, and physical, emotional, or sexual abuse and higher peak symptoms and higher peak velocity appeared stronger in females than males (**Table S5**). Further, the association between lower maternal education and older age at peak symptoms was only found in females (**Table S5**). There was no statistical evidence of sex differences in associations of domestic violence and psychiatric disorder with any outcomes.

## DISCUSSION

We used data from over >8,000 participants in the ALSPAC cohort to compare two methods for smoothing and estimating the severity and greatest rate of change of depressive symptoms trajectories through adolescence and examined how early life risk factors for adult depression relates to adolescent symptoms peak outcomes.

While both methods produced similar results, FPCA appeared to outperform PLME in estimating trajectories in females and in recovering trajectory peaks in both males and females. We found clear sex differences indicating that females had more adverse trajectories than males, including higher and earlier occurring peaks. Lastly, early life risk factors were associated with higher peak symptoms and peak velocity, with some also associating with the age of these peaks, and with evidence of sex differences indicating stronger associations in females than males for some risk factors.

To the best of our knowledge, this is the first study to apply FPCA to depressive symptoms trajectories. Our findings showed that FPCA appeared to outperform

PLME for recovering peaks in symptoms trajectories. FPCA treats trajectory as a continuous process rather than treating measurements as isolated points(Gertheiss et al., 2024). This approach may allow FPCA to better capture the complex dynamics of symptom evolution, which may make it more robust for identifying precise age and intensity of trajectory features like adolescent depressive symptom peaks. That PLME models tended to over-smooth trajectories might explain its lower peak estimates compared with FPCA.

Consistent with established evidence of a sex divergence in adolescent depression risk (Salk et al., 2017), we found that females had higher and steeper depressive symptoms trajectories, with higher peak symptoms and peak velocity, and that these peaks occurred earlier than in males. The sex differences reported here are also mostly consistent with earlier analyses of the ALSPAC cohort that used LME models with simpler global cubic polynomials, though that study did not report estimates for peak velocity (Kwong et al., 2019a). The earlier and accelerated symptoms development in females may be attributable to the, on average, earlier onset of puberty in females compared with males where rapid biological changes converge with concurrent psychosocial stressors to drive rapid symptom development (Hyde et al., 2008, Joinson et al., 2012, Tarif et al., 2025)

Our study reports novel associations of maternal education and perinatal depression and childhood traumas and psychiatric disorder with adolescent depressive symptoms rate of change and severity, which corroborate and build on previously reported associations with mental health in adolescents and young adults (Joinson et al., 2017, Tirumalaraju et al., 2020, Mulraney et al., 2021, Copeland et al., 2018, Croft et al., 2019). The observations that all risk factors (with the exception of maternal education) associated with an earlier peak velocity of depression symptoms aligns with the dimensional model of adversity and stress acceleration hypotheses (Colich et al., 2020, McLaughlin et al., 2014). Our finding that lower maternal education associated with older age at peak symptoms may be explained by the hypothesis that, compared with threat-related adversities, deprivation influences depression development through distinct neurocognitive mechanisms that may shift peak depressive vulnerability to later in adolescence (McLaughlin et al., 2014). SEP is a complex and multidimensional social construct and future studies using multiple indicators of SEP (and examining mediators) may shed light on mechanisms (Joinson et al., 2017). Finally, that some risk factor associations were stronger in females may be explained by adolescent females having a higher reactivity to interpersonal stressors compared to males (Rose and Rudolph, 2006, Morken et al., 2023).

Future work could examine approaches for directly modelling trajectory derivatives in FPCA and PLME rather than estimating these from the fitted trajectories (Liu and Müller, 2009, Dai et al., 2018, Hernández et al., 2023, Simpkin et al., 2018). Future studies could also explore the feasibility of single step modelling procedures for relating risk factors to trajectory peak outcomes to account for uncertainty in peak estimation (Yang et al., 2024, Zhang et al., 2023, Sayers et al., 2017). Investigating the suitability of FPCA for describing other mental health measures such as behavioural, emotional, and well-being trajectories (Armitage et al., 2023, Newland et al., 2019), including in different and combined cohorts is also warranted. Lastly, it is worth acknowledging that latent trajectory models are among the most popular methods of analysing depressive symptoms trajectories (Gataviņš et al., 2025, Weavers et al., 2023, Davies et al., 2020, Kwong et al., 2019b). However, these models aim to identify population subgroup trajectories and cannot be easily used to identify individual-specific peaks (Herle et al., 2020, Elhakeem et al., 2022).

Strengths of this study include the use of rich longitudinal data with a validated instrument for longitudinally assessing depressive symptoms, comparing two methods for estimating symptom patterns and their features and providing novel insights into how early life factors might associate with symptoms features in adolescence. Our trajectory analysis may have reduced selection bias due to missing SMFQ data by including all participants with two or more out of the ten SMFQ measurements under the missing at random assumption (i.e., the probability that an outcome value (SMFQ score) is missing depends on observed values of the outcome) (Fitzmaurice et al., 2009, Ségalas et al., 2024). Our trajectory analysis sample differed from those excluded due to missing data which can limit generalisability. Our early life risk factor association analysis was exploratory for the purpose of validating trajectory features and a more detailed study with full confounder adjustments and triangulation is required to examine the causal nature of the associations. These associations were also examined as part of two stage process and so does not take account of the uncertainty in our peak estimates; joint modelling approaches is an active area of research and would be useful (if feasible). Lastly, a lack of SMFQ measurement invariance across age and sex can hinder the consistency of depressive estimates however, analysis in ALSPAC supports the use of SMFQ for assessing symptoms trajectories across sex and the ages examined here (Schlechter et al., 2023).

In conclusion, our study extends the application of FPCA to depressive symptom trajectories and demonstrated its usefulness for identifying trajectory peak characteristics. We found that females exhibited higher and earlier peaks in both symptom severity and velocity, and that early life adversity was associated with the magnitude and timing of these peaks. If causality is established in future research, our findings would indicate that childhood stressors shift the underlying ‘tempo’ of symptom development to accelerate the timeline of clinical depression, highlighting children exposed to early life adversity as a priority target group for preventative interventions. Broadly, this work highlights the value of longitudinal population studies and the appropriate analysis of repeated measurements for improving our understanding of developmental mental health trajectories and their early determinants. Lastly, these methods could be used to model trajectory peaks not just in early life but across the lifespan in response to acute, macro-level stressors such as pandemics and war.

## Data availability

The informed consent obtained from ALSPAC (Avon Longitudinal Study of Parents and Children) participants does not allow the data to be made available through any third party maintained public repository. Supporting data are available from ALSPAC on request under the approved proposal number, B5149 Full instructions for applying for data access can be found here: http://www.bristol.ac.uk/alspac/researchers/access/. The ALSPAC study website contains details of all available data (http://www.bristol.ac.uk/alspac/researchers/our-data/). All analysis code used for this paper is made publicly available on a GitHub repository at https://github.com/LongitudinalModelling/smfq.

## Acknowledgements

We are extremely grateful to all of the families who took part in ALSPAC, the midwives for their help in recruiting them, and the whole ALSPAC team, which includes interviewers, computer and laboratory technicians, clerical workers, research scientists, volunteers, managers, receptionists and nurses.

## Ethical approval

Ethical approval for the study was obtained from the ALSPAC Ethics and Law Committee and the Local Research Ethics Committees (NHS Haydock REC: 10/H1010/70). Informed consent for the use of data was obtained from participants following the recommendations of the ALSPAC Ethics and Law Committee at the time. Participants can contact the study team at any time to retrospectively withdraw consent for their data to be used. Study participation is voluntary and during all data collection sweeps, information was provided on the intended use of data. At age 18 years, study children were sent ‘fair processing’ materials describing the intended use of their health and administrative records and given clear means to consent or object via a written form. Data were not extracted for participants who objected, or who were not sent fair processing materials.

