## Supplementary material for "Estimating severity and rate of change of depressive symptoms in adolescence: a comparison of functional principal component analysis and mixed effects models": Supplemantary_material

### Appendix S1. Supplementary methods

This appendix includes a description of the Short Mood and Feelings Questionnaire (SMFQ) items used to assess depressive symptoms and details on the derivation of the early life risk factors. It also describes the functional principal component analysis (FPCA) and P-spline linear mixed-effects (PLME) model used to analyse depressive symptoms trajectories and provides more details on the trajectory analysis and peak estimation.

***Depressive symptoms assessment with the SMFQ***

Depressive symptoms were self-reported between 2002-2018 across ten time points from mean age 10.6 to 25.8 years using the Short Mood and Feelings Questionnaire (SMFQ), which is a validated instrument for measuring depressive symptoms (Angold et al., 1995, Kwong, 2019, Turner et al., 2014, Eyre et al., 2021). At each timepoint, the presence of depressive symptoms during the previous two weeks was recorded by participant responses to the following 13 questions: “I felt miserable or unhappy”, “I didn’t enjoy anything at all”, “I felt so tired I just sat around and did nothing”, “I was very restless”, “I felt I was no good anymore”, “I cried a lot”, “I found it hard to think properly or concentrate”, “I hated myself”, “I was a bad person”, “I felt lonely”, “I thought nobody really loved me”, “I thought I could never be as good as others”, “I did everything wrong”. For each question, responses were reported on a 3-point scale (0 = not true, 1 = sometimes, 2 = true). Responses were summed to create a total score ranging from 0 to 26, with higher scores indicating greater depressive symptoms (Angold et al., 1995).

***Early life factors***

Maternal education

Early life socioeconomic position (SEP) was defined by maternal education (Galobardes et al., 2006). Maternal education was based on the mother’s highest educational attainment and was self-reported using questionnaires administered in pregnancy (32 weeks gestation). The responses were used to generate a categorical variable with increasing levels of achievement in the following five categories: none or certificate of secondary education (subject-specific qualifications of a lower level than ordinary-levels that were generally obtained at age 16 y (the minimal school leaving age from 1974 in England)), vocational, ordinary-level (subject-specific qualifications generally obtained at age 16 years), advanced-level (subject-specific qualifications generally obtained at age 18 years and which are required for university entry), and university degree. For this study, we used a harmonised maternal education variable that was derived from these five groups for the EU Child Cohort Network (Pinot de Moira et al., 2021). This variable was based on the International Standard Classification of Education 97 (ISCED-97) and consisted of 3 categories : low (no education to lower secondary; ISCED-97 categories 0–2), medium (upper and postsecondary; ISCED-97 categories 3–4), high (degree and above; ISCED-97 categories 5–6) (Cadman et al., 2023, Cadman et al., 2024, UNESCO United Nations Educational and Organization, 2003).

Perinatal maternal depression

Perinatal maternal depression was assessed using the Edinburgh Postnatal Depression Scale during pregnancy (at 18 and 32 weeks gestation) and 8 months postpartum (Paul and Pearson, 2020, Hermans et al., 2026). The Edinburgh Postnatal Depression Scale (Cox et al., 1987) is the most widely used perinatal depression instrument and consists of 10 items that assess depressive symptoms during the previous week.

Mothers rated the frequency of each symptom using four responses to the following 10 questions: “1. I have been able to laugh and see the funny side of things” (As much as I always could; Not quite so much now; Definitely not so much now; Not at all), “2. I have looked forward with enjoyment to things” (As much as I ever did; Rather less than I used to; Definitely less than I used to; Hardly at all), “3. I have blamed myself unnecessarily when things went wrong” (Yes, most of the time; Yes, some of the time; Not very often; No never), “4. I have been anxious or worried for no good reason” (No, not at all; Hardly ever; Yes, sometimes; Yes, often), “5. I have felt scared or panicky for no very good reason” (Yes, quite a lot; Yes, sometimes; No, not much; No, not at all), “6. Things have been getting on top of me” (Yes, most of the time; Yes, sometimes; No, hardly ever; No, not at all), “7. I have been so unhappy that I have had difficulty sleeping” (Yes, most of the time; Yes, sometimes; Not very often; No, not all), “8. I have felt sad or miserable” (Yes, most of the time; Yes, quite often; Not very often; No, not at all), “9. I have been so unhappy that I have been crying” (Yes, most of the time; Yes, quite often; Only occasionally; No, never), “10. The thought of harming myself has occurred to me” (Yes, quite often; Sometimes; Hardly ever; Never).

Responses are coded 0-3, with three items (1, 2, and 4) reverse coded, giving summary score ranging from 0-30, with higher indicating more depressive symptoms. We generated a binary variable to compare women with any perinatal depression (a score >12 for either prenatal or postnatal assessment) with those without perinatal depression (Paul and Pearson, 2020, Hermans et al., 2026).

Childhood traumas

Childhood trauma occurring before age 11 (0–10 years) was assessed prospectively using both parent and participant reports and were supplemented by participant reports at age 22 years about traumatic exposures that occurred in childhood (before 11 years). (Croft et al., 2019). The questions used to inform each trauma type exposure and responses regarding the severity and frequency of trauma exposure were carefully selected to ensure that a coding of exposed coding reflected an occurrence likely to be highly upsetting to anyone experiencing it. For this study, these responses were used to derive three binary variables: childhood domestic violence; a combined measure of physical, emotional, or sexual abuse; and bullying victimization. Detailed coding procedures for these specific items are described extensively elsewhere (Croft et al., 2019).

Exposure to domestic violence between ages 0 and 10 was evaluated using 15 questions. Questions focused on regular, physical acts of violence within the home that would be inherently traumatic for a child to witness. For questions addressing general violence between caregivers, any positive response was recorded as an indicator of domestic violence. For questions that referred to a specific instance or act of violence, for example ‘has your partner ever physically twisted your arm?’, only responses that referred to regular occurrences of this, as opposed to a single instance, were recorded as instances of domestic violence.

The derived variable for abuse consisted of physical, emotional, and sexual abuse. Physical abuse between ages 0 and 10 was identified using 32 parent questions and 5 participant questions, the latter being asked at age 22. Both mothers and their partners were asked whether they or their partner were physically cruel to their children and participants were asked if they had been physically hurt by their parents or another adult. A positive response to any of these questions was recorded as physical abuse. Emotional abuse was derived from 31 parent questions and 4 participant questions, three of which were asked at age 22. Parents were asked whether their children had been exposed to emotional cruelty by themselves or their partner, with any positive to response recorded as emotional abuse. Participants were asked whether adults had said hurtful or insulting things to them or if they were threatened with physical harm: responses of ‘often’ and ‘very often’ were recorded as an indicator of emotional abuse. Sexual abuse was identified using 7 questions from the parents and 4 questions from the participants, with 2 of the particpant questions asked at age 22. Caregivers’ reports of whether their child had been exposed to sexual abuse were recorded as indicators of sexual abuse in early life. Any positive response to questions that refer to any adult or older child forcing or attempting to force the participant into sexual activity was recorded as exposure. to sexual abuse. Questions that refer to partners pressuring the participants into sexual activity that referred to frequency of this occurring were recorded as indicators of sexual abuse if the participant responded ‘often’ or ‘all the time’. For this study, the three binary abuse variables were used to a binary variable for any physical, emotional or sexual abuse versus none.

Bullying victimisation between 0-10 years was assessed using 5 questions from the parents and 5 questions from the participants. Questions were selected from a comprehensive assessment of bullying that included a wide range of forms of bullying (i.e., name-calling, blackmail, assault). Bullying that would be the most likely to be highly distressing and traumatic was included: this included questioned that referred to any form of physical assault and threats of assault or blackmail. To be classified as a traumatic exposure, specific bullying instances must have occurred a minimum of four times within the past six months. Questions from caregivers and participants referring to bullying were used as indicators of bullying. For Questions that asked whether the child was being bullied, the response ‘certainly true’ was recorded as exposure to bullying.

Childhood psychiatric disorder

Childhood psychiatric disorders were assessed using parent-reported Development and Well-Being Assessment (DWABA) when the child was aged age 8 years (Goodman et al., 2000). DWABA consists of a package of questionnaires that are aimed at establishing the presence of common emotional, behavioural, and hyperactivity disorders in children and designed to generate ICD-10 and DSM-IV psychiatric diagnoses and is validated for epidemiological studies in the UK. The DWABA responses were used to derive DSM-IV diagnoses for attention deficit hyperactivity disorder (ADHD), oppositional or conduct disorder, pervasive developmental disorder, anxiety disorder, and depressive disorder. For this study, we created a binary variable to compare children any psychiatric disorder (i.e., any ADHD, any oppositional or conduct disorder, any pervasive developmental disorder, any anxiety disorder, or any depressive disorder) at age 8 years with those without a diagnosis.

***Functional Principal Component Analysis (FPCA)***

FPCA is a dimension reduction tool within Functional Data Analysis. It is founded upon the Karhunen-Loève expansion, which expresses each individual’s smooth trajectory as a population mean function and a weighted combination of the first K functional components that capture the main patterns of variability. The individual trajectory is therefore defined as:

$$y_{ij}=\mu\left( t_{ij} \right)+\sum_{k=1}^{K} {\xi_{ik}\phi}_{k}\left( t_{ij} \right)+\varepsilon_{ij}$$

(1)

where $y_{ij}$ denotes a single continuous repeated outcome (e.g., SMFQ score) measured in the i-th individual $\left( i=1, 2, \ldots, N \right)$ at age $t_{ij} \left( j=1, 2, \ldots, J_{i} \right)$, with the responses $y_{ij}$ assumed to be independent between individuals. $\mu\left( t_{ij} \right)$ is the population-average (mean) trajectory function. $\sum_{k=1}^{K} {\xi_{ik}\phi}_{k}\left( t_{ij} \right)$ captures the individual-specific deviation from the mean function, where $\phi_{k}\left( t_{ij} \right)$ are the orthonormal eigenfunctions (functional principal components), and $\xi_{ik}$ are the random functional principal component scores for the i-th individual, assumed to be uncorrelated across individuals and components. The independent measurement error $\varepsilon_{ij}$ assumed to have mean zero and variance.

For sparse or irregularly sampled longitudinal data, principal components analysis is carried out by smoothing the covariance function G(s,t) and using the conditional expectation (PACE) approach, which predicts the scores as conditional expectations given the observed data for each individual. However, because the classical PACE estimation can be computationally intensive as observations increase (Yao et al., 2005), we employed the Fast Covariance Estimation (FACE) algorithm proposed by Xiao et al. (2018). FACE serves as a computationally efficient implementation of FPCA by utilizing P-splines for smoothing mean and covariance functions, ensuring robustness even with irregular and sparse measurements.

Specifically, FACE estimates the covariance function by modelling it as a smooth surface over time using tensor-product B-splines,

$$G(s,t)\approx B\left( s \right)^{T}\Theta B\left( t \right) (2)$$

where $B(t)$ denote a B-spline basis and $\Theta$is a symmetric coefficient matrix estimated via penalised weighted least squares. The penalty term is equivalent to that used in bivariate P-splines (Eilers and Marx, 2003) and controls the smoothness of the estimated covariance surface. To ensure optimal performance, the smoothing parameter is selected using a fast leave-one-subject-out cross-validation procedure (see Xiao et al., 2018 for further methodological and computational details). Furthermore, this method employs a joint estimation approach for both the covariance function and the error variance, which helps distinguish the underlying functional trajectory to be separated from measurement error.

Once the smooth covariance surface $G(s,t)$ is estimated, an eigen-decomposition is performed to obtain the functional principal components (eigenfunctions) and their corresponding eigenvalues. Finally, the individual scores $\xi_{ik}$ are calculated, and individual trajectories are reconstructed using the first $K$ principal components, as shown in Equation (2). This methodology is implemented in the face package for R.

***P-spline Linear Mixed Effects model (PLME)***

Mixed effects models, also known as random effects or multilevel models, are widely used for analysing repeated measures data. These models combine two key components: fixed effects and random effects. Fixed effects describe the population-average (mean) trend across all individuals, while random effects capture between-individual variability by allowing subject-specific deviations from this overall trend. By incorporating both, mixed-effects models account for the correlation among repeated observations within the same individual.

One common approach is the linear mixed effects model (LME), which models the outcome as a linear combination of the fixed and random effects(Laird and Ware, 1982, Fitzmaurice et al., 2009). An LME model for a single continuous repeated outcome (e.g., SMFQ score) as a linear function of a single continuous predictor (e.g., age or time) that includes random intercepts and random slopes, is written as follows:

| $y_{ij}={(\beta}_{0}+ u_{0i})+{(\beta}_{1}+ u_{1i})\times t_{ij}+\varepsilon_{ij}$  ${(u}_{0i},u_{1i})^{t} \sim N\left( 0,\Omega_{u} \right); \varepsilon_{ij} \sim N\left( 0, \sigma_{\varepsilon}^{2} \right)$ | (3)  (3.1) |
| --- | --- |

where $y_{ij}$ denotes a single outcome measured in the i-th individual $\left( i=1, 2, \ldots, N \right)$ at age $t_{ij} \left( j=1, 2, \ldots, J_{i} \right)$, with the responses $y_{ij}$ assumed to be independent between individuals. The parameters $\beta_{o}$ and $\beta_{1}$ are fixed effects that represent the average intercept and average slope across all individuals in the population, respectively. The terms $u_{oi}$ and $u_{1i}$ are random effects that quantify how much the intercept and slope for the i-th individual deviate from the population-average intercept and slope, respectively. The random effects are assumed to be normally distributed with mean zero and covariance matrix $\Omega_{u}$. This assumption ensures that the fixed effects represent the population-level average. The residual error $\varepsilon_{ij}$ is assumed to be independently and identically normally distributed with mean zero and variance $\sigma_{\varepsilon}^{2}$. Random effects and residual errors are assumed to be mutually independent.

To extend this model to capture a nonlinear relationship between the outcome and age, we define the cubic P-spline linear mixed effect (PLME) model following the approach described by Djeundje and Currie (Djeundje and Currie, 2010). For the i-th subject at the j-th measurement time$t_{ij}$, the model is as follows:

|  | $y_{ij}=f\left( t_{ij} \right)+g_{i}\left( t_{ij} \right)+\varepsilon_{ij}$  $f(t_{ij})=\sum_{k=1}^{m} {\check{B}_{k}(t}_{ij};3){\alphǎ}_{k}$  $g_{i}(t_{ij})=\sum_{k=1}^{\tilde{m}} {\tilde{B}_{k}(t}_{ij};3)\tilde{\alpha}_{ki}$ | (4)  (4.1)  (4.2) |
| --- | --- | --- |

Where $\left\{ \check{B}_{k}(\cdot;3) :1 \leq k \leq m) \right\}$ is a set of $m$ cubic B-spline basis functions representing the population function $f$ (i.e., the fixed effects); $\left\{ \tilde{B}_{k}(\cdot;3) :1 \leq k \leq\tilde{m}) \right\}$ is a set of $\tilde{m}$ cubic B-spline basis functions representing the individual-specific deviations $g_{i}$ (i.e., random effects); and $\alphǎ={({\alphǎ}_{1},\ldots,{\alphǎ}_{m})}^{\top}$ and ${\tilde{\alpha}.}_{i}={(\tilde{\alpha}_{1i},\ldots,\tilde{\alpha}_{\tilde{m}i})}^{\top}$ are two coefficient vectors for the fixed and random B-splines, respectively. To complete the full P-spline specification of the model in Eq. (4), we define two $p$-th order difference penalties for $f$ and $g_{i}$ as:

| $\check{P}_{\check{\lambda}=\check{\lambda}_{1}}={{\check{\lambda}_{1}\check{D}}_{p}}^{\top}\check{D}_{p}$ | (5.1) |
| --- | --- |
| $\tilde{P}_{\tilde{\lambda}=\left( \tilde{\lambda}_{0},\tilde{\lambda}_{1} \right)}=\tilde{\lambda}_{0}I$+$\tilde{\lambda}_{1}{\tilde{D}_{p}}^{\top}\tilde{D}_{p}$ | (5.2) |

Following the work of Eilers and Marx (1996) (Eilers and Marx, 1996), we employ discrete penalties where $\check{D}_{p}$ and $\tilde{D}_{p}$ are constructed as $p$-th order difference matrices to discourage large jumps between adjacent basis coefficients. A common choice for enforcing smoothness is the second-order difference ($p=2$). Allowing different values of $p$ for each smooth term in the model e.g., using $p=2$ for $f$ and $p=1$ for $g_{i}$, can help reduce collinearity between the population-level and individual-level smoothers(Pedersen et al., 2019, Baayen et al., 2018). $\check{\lambda}_{1}$ and $\tilde{\lambda}_{1}$ are two smoothing parameters that determine the strength of the penalty on the fixed ($\alphǎ$) and random (${\tilde{\alpha}.}_{i}$) basis coefficients, respectively. $\tilde{\lambda}_{0}$ has an additional ridge penalty $I$ that addresses identifiability issues (Djeundje and Currie, 2010).

***Further details on trajectory analysis and estimation of peak symptoms and peak velocity in ALSPAC***

For both FPCA and PLME approaches, P-splines were constructed using a cubic B-splines (De Boor, 1978). We compared models with 5, 6, and 7 equally spaced knots, and 6 knots were selected based on the lowest root mean square error (RMSE). For PLME, a model with 7 knots in the fixed spline (and 6 in the random spline) was used. We applied a 2^nd^ order difference penalty for the P-spline in FPCA and for the fixed effect P-spline in PLME. A 1^st^ order difference penalty was used for the random P-spline (in PLME) to avoid collinearity with the fixed effect P-spline.

The magnitude and age of peak depressive symptoms during adolescence was estimated using the second derivative of the predicted trajectories up to age 20 years. We first identified the approximate location of a turning point that indicated a peak by calculating where the smoothed second derivative changes sign from positive to negative. The location of this turning point was then refined by iteratively fitting a quadratic model to an expanding set of neighbouring data points until a well-defined parabolic curve was established. The magnitude (SMFQ score) and age (years) of peak depressive symptoms was calculated from the vertex of this final parabola. The same approach was then repeated on the predicted velocity curves up to age 20 years (i.e., the first derivative of the predicted trajectories) to estimate the peak (score/year) and age at peak depressive symptoms velocity. Symptom velocity curves were obtained using cubic smoothing spline differentiation of each individual's fitted trajectory at each point on the age grid and the same sign change procedure was then used to identify peak symptoms velocity (rate of most rapid increase in depressive symptoms: score/year) and the corresponding age at peak velocity (years). Peak feature extraction was implemented using the getPeak function from the sitar package in R, with the Dy = TRUE argument used for velocity-based features.

***References***

ANGOLD, A., COSTELLO, E. J., MESSER, S. C. & PICKLES, A. (1995). Development of a short questionnaire for use in epidemiological studies of depression in children and adolescents. International Journal of Methods in Psychiatric Research, 5, 237-249.

BAAYEN, R. H., VAN RIJ, J., DE CAT, C. & WOOD, S. (2018). Autocorrelated Errors in Experimental Data in the Language Sciences: Some Solutions Offered by Generalized Additive Mixed Models. In D. SPEELMAN, K. HEYLEN & D. GEERAERTS (Eds.) Mixed-Effects Regression Models in Linguistics. (pp. 49-69). Cham: Springer International Publishing.

CADMAN, T., AVRAAM, D., CARSON, J., ELHAKEEM, A., GROTE, V., GUERLICH, K., GUXENS, M., HOWE, L. D., HUANG, R.-C., HARRIS, J. R., HOUWELING, T. A. J., HYDE, E., JADDOE, V., JANSEN, P. W., JULVEZ, J., KOLETZKO, B., LIN, A., MARGETAKI, K., MELCHIOR, M., NADER, J. T., PEDERSEN, M., PIZZI, C., ROUMELIOTAKI, T., SWERTZ, M., TAFFLET, M., TAYLOR-ROBINSON, D., WOOTTON, R. E. & STRANDBERG-LARSEN, K. (2024). Social inequalities in child mental health trajectories: a longitudinal study using birth cohort data 12 countries. BMC Public Health, 24, 2930.

CADMAN, T., ELHAKEEM, A., VINTHER, J. L., AVRAAM, D., CARRASCO, P., CALAS, L., CARDO, M., CHARLES, M. A., CORPELEIJN, E., CROZIER, S., DE CASTRO, M., ESTARLICH, M., FERNANDES, A., FOSSATTI, S., GRUSZFELD, D., GURLICH, K., GROTE, V., HAAKMA, S., HARRIS, J. R., HEUDE, B., HUANG, R. C., IBARLUZEA, J., INSKIP, H., JADDOE, V., KOLETZKO, B., LUQUE, V., MANIOS, Y., MOIRANO, G., MOSCHONIS, G., NADER, J., NIEUWENHUIJSEN, M., ANDERSEN, A. N., MCEACHEN, R., DE MOIRA, A. P., POPOVIC, M., ROUMELIOTAKI, T., SALIKA, T., MARINA, L. S., SANTOS, S., SERBERT, S., TZOROVILI, E., VAFEIADI, M., VERDUCI, E., VRIJHEID, M., VRIJKOTTE, T. G. M., WELTEN, M., WRIGHT, J., YANG, T. C., ZUGNA, D. & LAWLOR, D. (2023). Associations of Maternal Educational Level, Proximity to Greenspace During Pregnancy, and Gestational Diabetes With Body Mass Index From Infancy to Early Adulthood: A Proof-of-Concept Federated Analysis in 18 Birth Cohorts. Am J Epidemiol.

COX, J. L., HOLDEN, J. M. & SAGOVSKY, R. (1987). Detection of postnatal depression. Development of the 10-item Edinburgh Postnatal Depression Scale. Br J Psychiatry, 150, 782-786.

CROFT, J., HERON, J., TEUFEL, C., CANNON, M., WOLKE, D., THOMPSON, A., HOUTEPEN, L. & ZAMMIT, S. (2019). Association of Trauma Type, Age of Exposure, and Frequency in Childhood and Adolescence With Psychotic Experiences in Early Adulthood. JAMA Psychiatry, 76, 79-86.

DE BOOR, C. (1978). A practical guide to splines, New York: Springer-Verlag.

DJEUNDJE, V. A. B. & CURRIE, I. D. (2010). Appropriate covariance-specification via penalties for penalized splines in mixed models for longitudinal data. Electronic Journal of Statistics, 4, 1202-1224, 1223.

EILERS, P. H. C. & MARX, B. D. (1996). Flexible smoothing with <i>B</i>-splines and penalties. Statistical Science, 11, 89-121, 133.

EILERS, P. H. C. & MARX, B. D. (2003). Multivariate calibration with temperature interaction using two-dimensional penalized signal regression. Chemometrics and Intelligent Laboratory Systems, 66, 159-174.

EYRE, O., BEVAN JONES, R., AGHA, S. S., WOOTTON, R. E., THAPAR, A. K., STERGIAKOULI, E., LANGLEY, K., COLLISHAW, S., THAPAR, A. & RIGLIN, L. (2021). Validation of the short Mood and Feelings Questionnaire in young adulthood. J Affect Disord, 294, 883-888.

FITZMAURICE, G., DAVIDIAN, M., VERBEKE, G. & MOLENBERGHS, G. (2009). Longitudinal data analysis, USA: Chapman & Hall/CRC.

GALOBARDES, B., SHAW, M., LAWLOR, D. A., LYNCH, J. W. & DAVEY SMITH, G. (2006). Indicators of socioeconomic position (part 1). J Epidemiol Community Health, 60, 7-12.

GOODMAN, R., FORD, T., RICHARDS, H., GATWARD, R. & MELTZER, H. (2000). The Development and Well-Being Assessment: description and initial validation of an integrated assessment of child and adolescent psychopathology. J Child Psychol Psychiatry, 41, 645-655.

HERMANS, A. P. C., AVRAAM, D., SCHUURMANS, I. K., SOARES, A. G., LAHTI-PULKKINEN, M., GIRCHENKO, P., VRIJKOTTE, T. G. M., DE ROOIJ, S. R., ELHAKEEM, A., VAN DER WAERDEN, J., HEUDE, B., VAINQUEUR, C., YANG, T. C., CHEUNG, R. W., LEWER, D., STRANDBERG-LARSEN, K., CADMAN, T., POPOVIC, M., CANDELORA, F., LAHTI, J., RÄIKKÖNEN, K., CECIL, C. A. M. & EL MARROUN, H. (2026). Prenatal maternal depression and child behavioural and developmental outcomes: an individual participant data meta-analysis in 76,514 children from the EU Child Cohort Network. The Lancet Regional Health – Europe, 63.

KWONG, A. (2019). Examining the longitudinal nature of depressive symptoms in the Avon Longitudinal Study of Parents and Children (ALSPAC). Wellcome Open Res, 4, 126.

LAIRD, N. M. & WARE, J. H. (1982). Random-effects models for longitudinal data. Biometrics, 38, 963-974.

PAUL, E. & PEARSON, R. M. (2020). Depressive symptoms measured using the Edinburgh Postnatal Depression Scale in mothers and partners in the ALSPAC Study: A data note. Wellcome Open Res, 5, 108.

PEDERSEN, E. J., MILLER, D. L., SIMPSON, G. L. & ROSS, N. (2019). Hierarchical generalized additive models in ecology: an introduction with mgcv. PeerJ, 7, e6876-e6876.

PINOT DE MOIRA, A., HAAKMA, S., STRANDBERG-LARSEN, K., VAN ENCKEVORT, E., KOOIJMAN, M., CADMAN, T., CARDOL, M., CORPELEIJN, E., CROZIER, S. & ELHAKEEM, A. (2021). The EU Child Cohort Network’s core data: establishing a set of findable, accessible, interoperable and re-usable (FAIR) variables. European Journal of Epidemiology.

TURNER, N., JOINSON, C., PETERS, T. J., WILES, N. & LEWIS, G. (2014). Validity of the Short Mood and Feelings Questionnaire in late adolescence. Psychol Assess, 26, 752-762.

UNESCO UNITED NATIONS EDUCATIONAL, S. & ORGANIZATION, C. (2003). International standard classification of education, ISCED 1997. Advances in Cross-National Comparison: A European Working Book for Demographic and Socio-Economic Variables. (pp. 195-220). Springer.

YAO, F., MÜLLER, H.-G. & WANG, J.-L. (2005). Functional Data Analysis for Sparse Longitudinal Data. Journal of the American Statistical Association, 100, 577-590.

### Table S1. Comparison of participants included in the depressive symptoms score trajectory analysis with those that were excluded due to having fewer than two or no SMFQ measures.

|  | Included in trajectory analysis, n=8,264 | Excluded from trajectory analysis, n=6,692 |
| --- | --- | --- |
| Sex, No. (%) |  |  |
| *males* | 3800 (46.0) | 3817 (57.0) |
| *females* | 4464 (54.0) | 2875 (43.0) |
| Maternal education |  |  |
| *High* | 1221 (16.4) | 373 (7.6) |
| *Medium* | 5202 (70.0) | 3064 (62.2) |
| *Low* | 1008 (13.6) | 1487 (30.2) |
| Prenatal maternal depression |  |  |
| *No* | 5623 (82.4) | 2989 (71.5) |
| *Yes* | 1202 (17.6) | 1191 (28.5) |
| Domestic violence |  |  |
| *No* | 5871 (80.4) | 2302 (70.3) |
| *Yes* | 1435 (19.6) | 973 (29.7) |
| Physical, emotional, or sexual abuse |  |  |
| *No* | 5480 (75.4) | 2210 (76.4) |
| *Yes* | 1789 (24.6) | 683 (23.6) |
| Bullying victimisation |  |  |
| *No* | 5290 (77.6) | 1755 (82.9) |
| *Yes* | 1528 (22.4) | 361 (17.1) |
| Psychiatric disorder |  |  |
| *No* | 6000 (93.5) | 1605 (90.1) |
| *Yes* | 418 (6.5) | 177 (9.9) |

### Table S2. Magnitude and age of peak depressive symptoms and peak symptoms velocity.

|  | Shared samples^a^ | | Unique samples^b^ | |
| --- | --- | --- | --- | --- |
|  | Males (n=2,232) | Females (n=3,905) | Males | Females |
| FPCA estimates |  |  |  |  |
| *Peak symptoms* | 5.4 (2.5) | 7.9 (3.8) | 5.8 (2.7) | 8.1 (3.9) |
| *Age at peak symptoms* | 18.0 (1.0) | 17.0 (1.8) | 18.1 (1.2) | 17.0 (1.8) |
| *Peak velocity* | 0.5 (0.4) | 1.3 (0.9) | 0.6 (0.5) | 1.4 (0.9) |
| *Age at peak velocity* | 16.0 (1.1) | 14.4 (1.8) | 16.1 (1.3) | 14.4 (1.8) |
| PLME estimates |  |  |  |  |
| *Peak symptoms* | 5.2 (2.0) | 7.2 (2.9) | 5.0 (2.0) | 7.2 (2.8) |
| *Age at peak symptoms* | 18.6 (0.8) | 17.6 (0.8) | 18.6 (0.8) | 17.6 (0.8) |
| *Peak velocity* | 0.3 (0.1) | 1.0 (0.2) | 0.4 (0.2) | 1.0 (0.2) |
| *Age at peak velocity* | 15.6 (0.4) | 13.5 (0.1) | 15.8 (0.4) | 13.5 (0.1) |

^a^ Shared samples are participants with estimates for magnitude and age of peak symptoms and peak and age at peak symptoms velocity from FPCA and PLME. ^b^ The numbers of participants with peak and age at peak symptoms were 3,226 males, 4,280 females (FPCA), and 2,476 males, 3,970 females (PLME). The numbers of participants with magnitude and age of peak symptoms velocity were 3,793 males, 4,461 females for both FPCA and PLME.

### Table S3. Pearson correlation coefficients between FPCA and PLME based estimates of the magnitude and age of peak depressive symptoms and peak symptoms velocity

|  | Males | Females |
| --- | --- | --- |
| A. Peak depressive symptoms and age at peak symptoms |  |  |
| *FPCA* | -0.15 | 0.03 |
| *PLME* | -0.06 | 0.13 |
| B. Peak symptoms velocity and age at peak velocity |  |  |
| *FPCA* | 0.02 | -0.02 |
| *PLME* | -0.18 | 0.28 |
| C. FPCA and PLME |  |  |
| *Peak depressive symptoms* | 0.97 | 0.95 |
| *Age at peak symptoms* | 0.52 | 0.41 |
| *Peak symptoms velocity* | 0.56 | 0.52 |
| *Age at peak velocity* | 0.47 | 0.33 |

Table shows Pearson correlations within each method between peak/age of peak depressive symptoms (A) and peak symptoms velocity (B), and Pearson correlations between pairs of FPCA and PLME based estimates of the magnitude and timing of peak depressive symptoms and peak symptoms velocity (C). All correlation coefficients were from 6,137 participants with all depressive symptoms estimates from both methods (2,232 males, 3,905 females).

### Table S4. Association of early life factors with FPCA, PLME and FPCA PLME average based depressive symptoms outcomes.

|  | Mean difference (95% CI) | | |
| --- | --- | --- | --- |
|  | FPCA  N=7,514 | LME  n=6,451 | FPCA-PLME  n=6,137 |
| ***Peak symptoms score*** |  |  |  |
| Sex |  |  |  |
| *Males* | 0 (ref) | 0 (ref) | 0 (ref) |
| *Females* | 2.36 (2.21 to 2.52) | 2.2 (2.07 to 2.33) | 2.19 (2.04 to 2.35) |
| Maternal education^a^ |  |  |  |
| *High* | 0 (ref) | 0 (ref) | 0 (ref) |
| *Medium* | 0.26 (0.03 to 0.49) | 0.31 (0.13 to 0.49) | 0.30 (0.09 to 0.52) |
| *Low* | 0.55 (0.25 to 0.85) | 0.57 (0.32 to 0.81) | 0.58 (0.30 to 0.87) |
| Maternal depression^b^ |  |  |  |
| *No* | 0 (ref) | 0 (ref) | 0 (ref) |
| *Yes* | 1.13 (0.90 to 1.36) | 1.05 (0.86 to 1.24) | 1.14 (0.92 to 1.36) |
| Domestic violence^b^ |  |  |  |
| *No* | 0 (ref) | 0 (ref) | 0 (ref) |
| *Yes* | 0.63 (0.42 to 0.85) | 0.64 (0.47 to 0.82) | 0.64 (0.44 to 0.84) |
| Physical, emotional, or sexual abuse^b^ |  |  |  |
| *No* | 0 (ref) | 0 (ref) | 0 (ref) |
| *Yes* | 1.28 (1.09 to 1.48) | 0.98 (0.82 to 1.14) | 1.07 (0.88 to 1.26) |
| Bullying victimisation^b^ |  |  |  |
| *No* | 0 (ref) | 0 (ref) | 0 (ref) |
| *Yes* | 1.19 (0.98 to 1.4) | 1.02 (0.85 to 1.19) | 1.13 (0.93 to 1.33) |
| Psychiatric disorder^b^ |  |  |  |
| *No* | 0 (ref) | 0 (ref) | 0 (ref) |
| *Yes* | 1.28 (0.90 to 1.66) | 0.99 (0.68 to 1.31) | 1.08 (0.71 to 1.46) |
| ***Age at peak symptoms*** |  |  |  |
| Sex |  |  |  |
| *Males* | 0 (ref) | 0 (ref) | 0 (ref) |
| *Females* | -1.1 (-1.17 to -1.02) | -1.04 (-1.08 to -1.00) | -1.06 (-1.11 to -1.01) |
| Maternal education^a^ |  |  |  |
| *High* | 0 (ref) | 0 (ref) | 0 (ref) |
| *Medium* | 0.17 (0.06 to 0.27) | 0.13 (0.07 to 0.18) | 0.17 (0.09 to 0.24) |
| *Low* | 0.31 (0.17 to 0.45) | 0.18 (0.10 to 0.25) | 0.26 (0.16 to 0.36) |
| Maternal depression^b^ |  |  |  |
| *No* | 0 (ref) | 0 (ref) | 0 (ref) |
| *Yes* | 0.04 (-0.07 to 0.15) | 0.03 (-0.03 to 0.09) | 0.02 (-0.06 to 0.10) |
| Domestic violence^b^ |  |  |  |
| *No* | 0 (ref) | 0 (ref) | 0 (ref) |
| *Yes* | -0.03 (-0.13 to 0.07) | 0.01 (-0.04 to 0.06) | -0.01 (-0.08 to 0.06) |
| Physical, emotional, or sexual abuse^b^ |  |  |  |
| *No* | 0 (ref) | 0 (ref) | 0 (ref) |
| *Yes* | -0.12 (-0.22 to -0.03) | -0.03 (-0.07 to 0.02) | -0.06 (-0.12 to 0.01) |
| Bullying victimisation^b^ |  |  |  |
| *No* | 0 (ref) | 0 (ref) | 0 (ref) |
| *Yes* | -0.23 (-0.33 to -0.13) | -0.12 (-0.17 to -0.07) | -0.16 (-0.23 to -0.09) |
| Psychiatric disorder^b^ |  |  |  |
| *No* | 0 (ref) | 0 (ref) | 0 (ref) |
| *Yes* | -0.08 (-0.26 to 0.10) | -0.02 (-0.12 to 0.08) | -0.04 (-0.17 to 0.09) |
| ***Peak symptoms velocity*** |  |  |  |
| Sex |  |  |  |
| *Males* | 0 (ref) | 0 (ref) | 0 (ref) |
| *Females* | 0.74 (0.70 to 0.77) | 0.63 (0.63 to 0.64) | 0.70 (0.68 to 0.72) |
| Maternal education^a^ |  |  |  |
| *High* | 0 (ref) | 0 (ref) | 0 (ref) |
| *Medium* | 0.06 (0.01 to 0.11) | 0.02 (0.01 to 0.03) | 0.04 (0.01 to 0.07) |
| *Low* | 0.09 (0.02 to 0.15) | 0.02 (0.01 to 0.04) | 0.05 (0.01 to 0.09) |
| Maternal depression^b^ |  |  |  |
| *No* | 0 (ref) | 0 (ref) | 0 (ref) |
| *Yes* | 0.14 (0.08 to 0.19) | 0.02 (0.00 to 0.03) | 0.08 (0.05 to 0.11) |
| Domestic violence^b^ |  |  |  |
| *No* | 0 (ref) | 0 (ref) | 0 (ref) |
| *Yes* | 0.04 (0.00 to 0.09) | 0.00 (-0.01 to 0.01) | 0.03 (0 to 0.06) |
| Physical, emotional, or sexual abuse^b^ |  |  |  |
| *No* | 0 (ref) | 0 (ref) | 0 (ref) |
| *Yes* | 0.19 (0.15 to 0.23) | 0.01 (0.00 to 0.02) | 0.10 (0.07 to 0.12) |
| Bullying victimisation^b^ |  |  |  |
| *No* | 0 (ref) | 0 (ref) | 0 (ref) |
| *Yes* | 0.14 (0.1 to 0.19) | -0.02 (-0.03 to -0.01) | 0.07 (0.04 to 0.1) |
| Psychiatric disorder^b^ |  |  |  |
| *No* | 0 (ref) | 0 (ref) | 0 (ref) |
| *Yes* | 0.13 (0.05 to 0.22) | 0.00 (-0.02 to 0.02) | 0.05 (0.00 to 0.10) |
| ***Age at peak velocity*** |  |  |  |
| Sex |  |  |  |
| *Males* | 0 (ref) | 0 (ref) | 0 (ref) |
| *Females* | -1.59 (-1.66 to -1.52) | -2.22 (-2.23 to -2.20) | -1.88 (-1.92 to -1.84) |
| Maternal education^a^ |  |  |  |
| *High* | 0 (ref) | 0 (ref) | 0 (ref) |
| *Medium* | 0.09 (-0.01 to 0.19) | 0.03 (0.01 to 0.04) | 0.05 (-0.01 to 0.11) |
| *Low* | 0.06 (-0.07 to 0.20) | 0.05 (0.02 to 0.07) | 0.04 (-0.04 to 0.12) |
| Maternal depression^b^ |  |  |  |
| *No* | 0 (ref) | 0 (ref) | 0 (ref) |
| *Yes* | -0.16 (-0.27 to -0.06) | -0.02 (-0.04 to 0.00) | -0.11 (-0.17 to -0.05) |
| Domestic violence^b^ |  |  |  |
| *No* | 0 (ref) | 0 (ref) | 0 (ref) |
| *Yes* | -0.18 (-0.27 to -0.08) | -0.01 (-0.03 to 0.01) | -0.08 (-0.13 to -0.02) |
| Physical, emotional, or sexual abuse^b^ |  |  |  |
| *No* | 0 (ref) | 0 (ref) | 0 (ref) |
| *Yes* | -0.23 (-0.32 to -0.14) | -0.03 (-0.05 to -0.01) | -0.13 (-0.18 to -0.07) |
| Bullying victimisation^b^ |  |  |  |
| *No* | 0 (ref) | 0 (ref) | 0 (ref) |
| *Yes* | -0.33 (-0.42 to -0.23) | -0.01 (-0.03 to 0.01) | -0.17 (-0.23 to -0.11) |
| Psychiatric disorder^b^ |  |  |  |
| *No* | 0 (ref) | 0 (ref) | 0 (ref) |
| *Yes* | -0.28 (-0.44 to -0.11) | 0 (-0.04 to 0.03) | -0.14 (-0.25 to -0.04) |

FPCA-PLME average calculated as the mean of the PLME and FPCA estimates.

^a^ adjusted for sex. ^b^ adjusted for sex and maternal education.

### Table S5. Association of early life factors with magnitude and age of peak depressive symptoms and peak symptoms velocity in males and females.

|  | Males | Females | P sex int |
| --- | --- | --- | --- |
| **Peak symptoms (score)** |  |  |  |
| Maternal education |  |  | 0.01 |
| *High* | 0 (ref) | 0 (ref) |  |
| *Medium* | -0.03 (-0.53 to 0.48) | 0.48 (0.03 to 0.92) |  |
| *Low* | 0.02 (-0.65 to 0.70) | 0.95 (0.36 to 1.53) |  |
| Maternal depression |  |  | 0.008 |
| *No* | 0 (ref) | 0 (ref) |  |
| *Yes* | 0.78 (0.32 to 1.24) | 1.40 (1.00 to 1.80) |  |
| Physical, emotional, or sexual abuse^b^ |  |  | 0.00002 |
| *No* | 0 (ref) | 0 (ref) |  |
| *Yes* | 0.79 (0.40 to 1.19) | 1.65 (1.31 to 1.99) |  |
| Bullying victimisation |  |  | 0.07 |
| *No* | 0 (ref) | 0 (ref) |  |
| *Yes* | 1 (0.61 to 1.38) | 1.4 (0.99 to 1.8) |  |
| **Age at peak symptoms (y)** |  |  |  |
| Maternal education |  |  | 0.07 |
| *High* | 0 (ref) | 0 (ref) |  |
| *Medium* | 0.04 (-0.20 to 0.27) | 0.27 (0.06 to 0.47) |  |
| *Low* | 0.14 (-0.18 to 0.45) | 0.44 (0.16 to 0.71) |  |
| **Peak velocity (score / y)** |  |  |  |
| Maternal education |  |  | 0.04 |
| *High* | 0 (ref) | 0 (ref) |  |
| *Medium* | 0.00 (-0.11 to 0.11) | 0.11 (0.01 to 0.20) |  |
| *Low* | -0.01 (-0.16 to 0.14) | 0.16 (0.03 to 0.29) |  |
| Maternal depression |  |  | 0.03 |
| *No* | 0 (ref) | 0 (ref) |  |
| *Yes* | 0.07 (-0.03 to 0.17) | 0.18 (0.10 to 0.27) |  |
| Physical, emotional, or sexual abuse |  |  | 0.0000006 |
| *No* | 0 (ref) | 0 (ref) |  |
| *Yes* | 0.06 (-0.02 to 0.15) | 0.29 (0.21 to 0.36) |  |
| Bullying victimisation |  |  | 0.0002 |
| *No* | 0 (ref) | 0 (ref) |  |
| *Yes* | 0.06 (-0.02 to 0.14) | 0.24 (0.15 to 0.33) |  |

Estimates from separate models with risk factor by sex interactions and adjusted for maternal education (for the other risk factor models).

### Figure S1. Predicted depressive symptoms velocity trajectories.

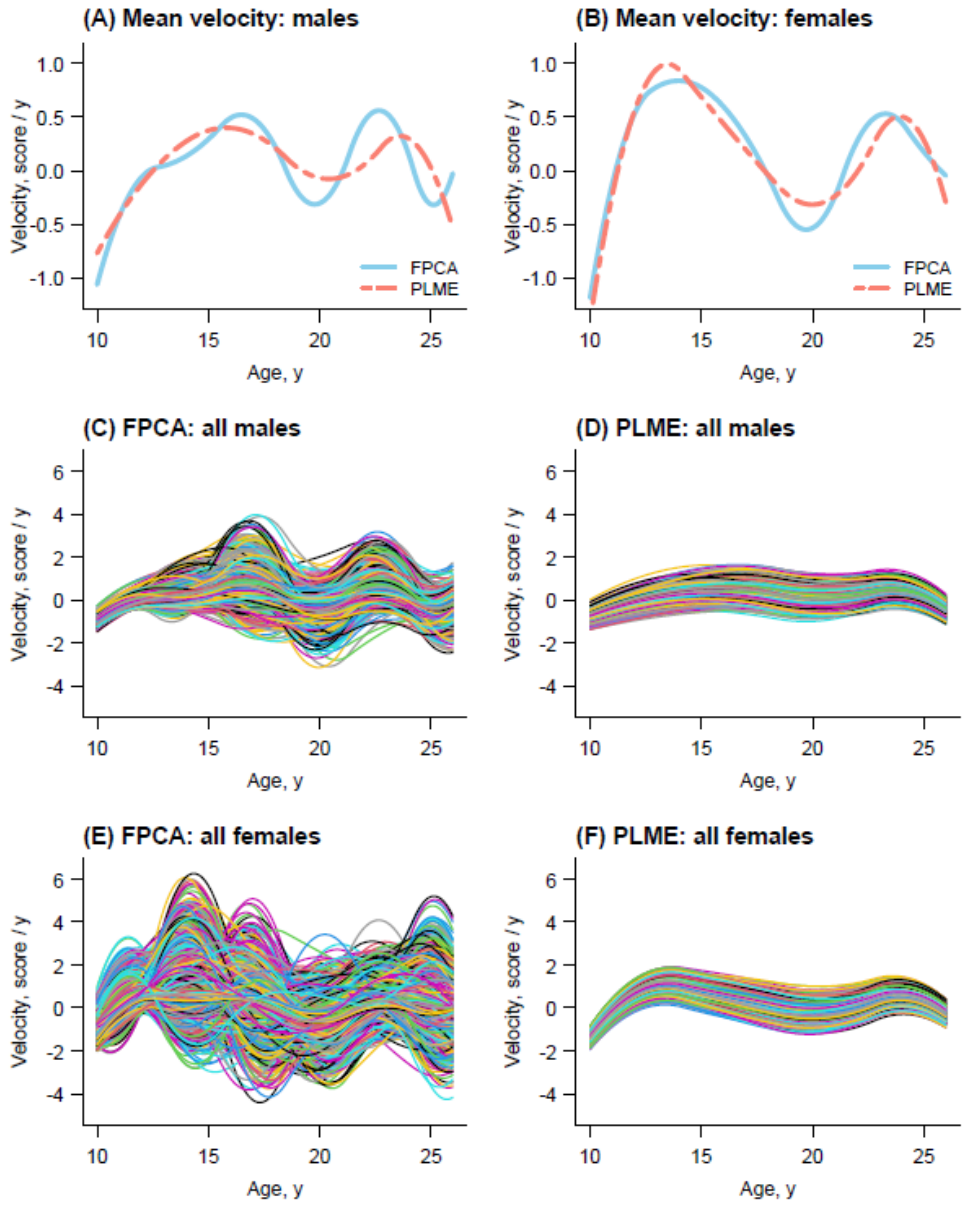

Figure shows the FPCA and PLME predicted population-average SMFQ depressive symptoms velocity trajectories in males and females (A, B), and the FPCA and PLME predicted individual-specific velocity trajectories in males (C, D) and females (E, F).

### Figure S2. FPCA and PLME model residuals against age

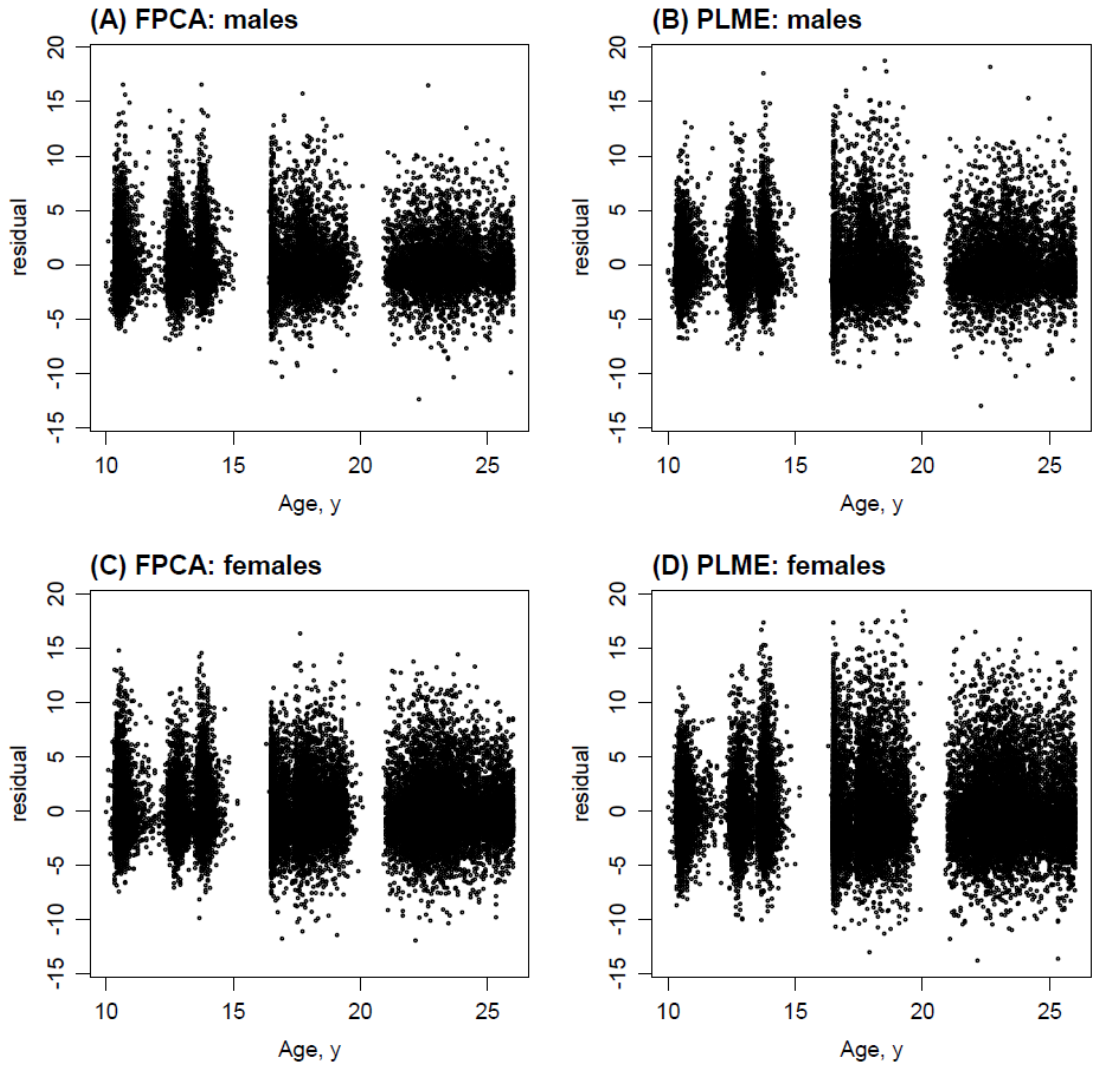

Figure shows the FPCA and PLME model residuals (observed SMFQ score - predicted SMFQ score) plotted against age in males (A, B) and females (C, D).

### Figure S3. Bland-Altman analysis of agreement between FPCA and PLME based estimates of the magnitude and age of peak depressive symptoms and peak symptoms velocity.

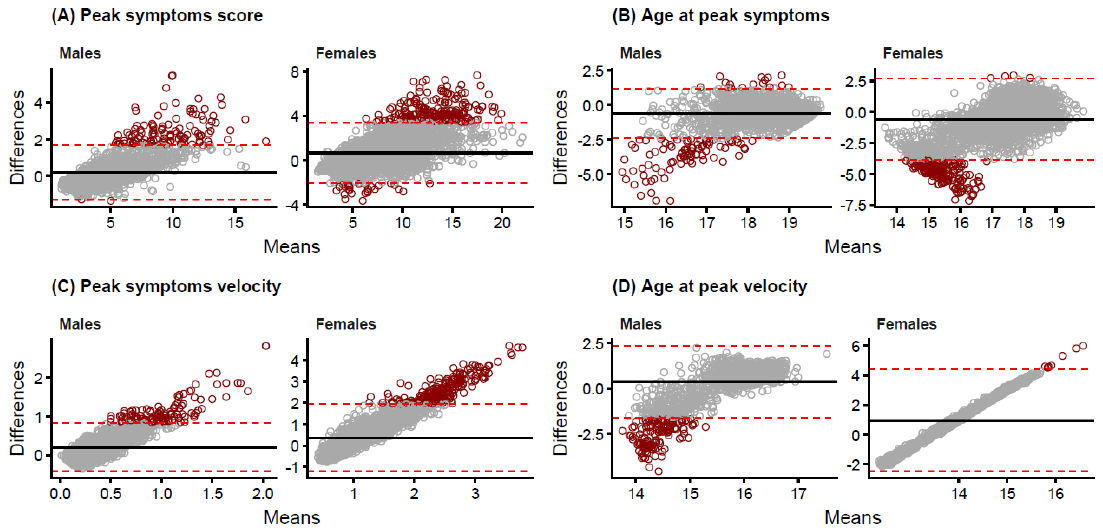

Figure shows plots of the difference between the FPCA and PLME based estimates (FPCA minus PLME) on the y-axis against the means of the two estimates for peak depressive symptoms score (A) and the age at peak symptoms (B), and for peak symptoms velocity (C) and the age at peak velocity (D), separately in males and females. The solid black line represents the mean difference between the two estimates. The dashed red lines represent the 95% limits of agreement (mean difference ± 1.96 standard deviations of the differences. The dark red dots represent estimates that fall outside of the 95% limits of agreement. All other estimates (within the limits) are shown in dark grey.

### Figure S4. Schematic representation of FPCA and PLME approaches for modelling longitudinal trajectories

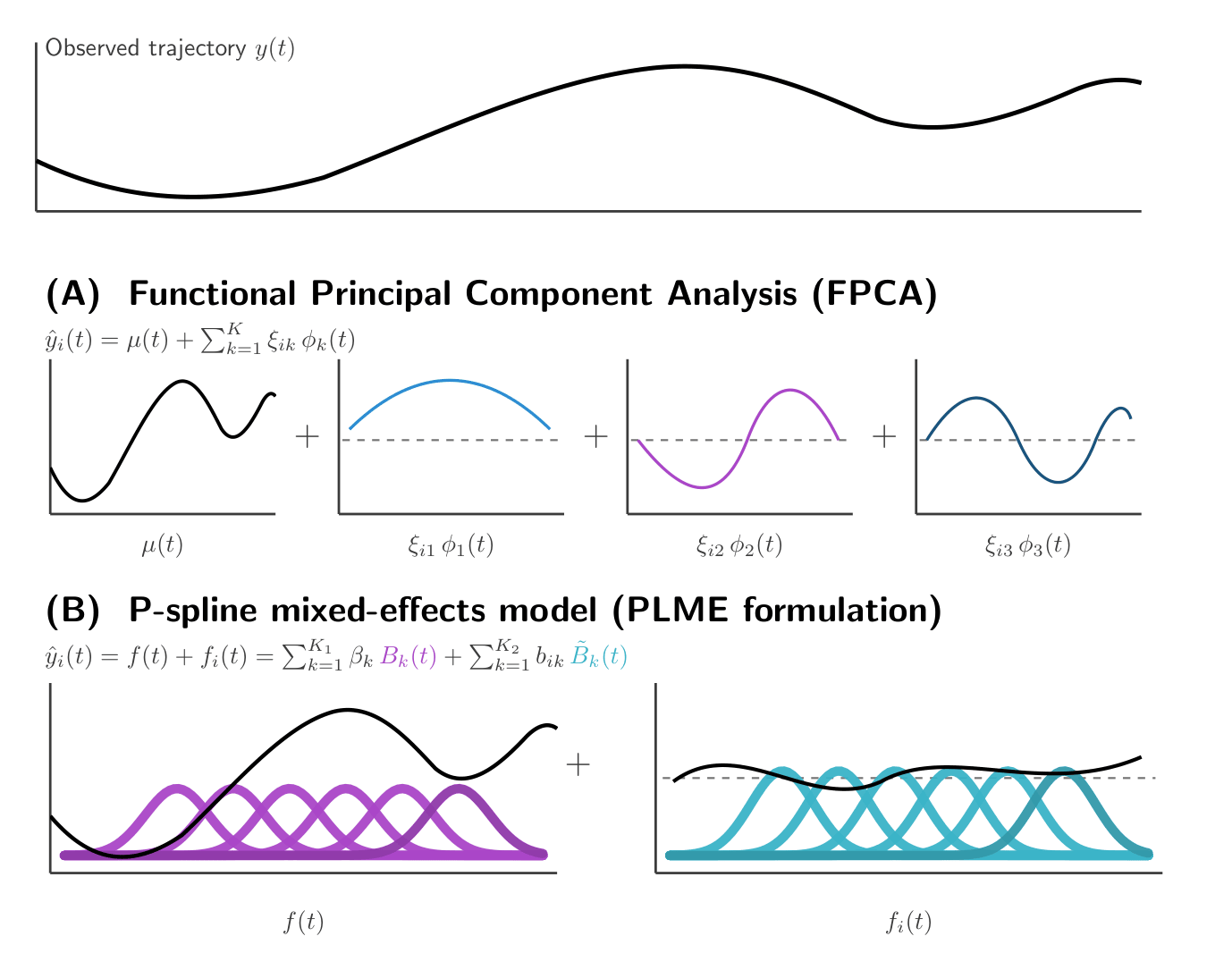

This Figure shows (A) FPCA: the individual trajectory$y_{i}\left( t \right)$ is decomposed into a population mean function$\mu$(t) and subject-specific deviations expressed as weighted FPC. (B) P-spline mixed-effects model (PLME formulation): the trajectory is represented as the sum of a smooth fixed effect $f\left( t \right)$ and an individual-specific deviation $f_{i}\left( t \right)$, both expressed as linear combinations of B-spline basis functions.
